# Stratified evaluation of blood RNA sequencing in a rare disease cohort

**DOI:** 10.64898/2026.05.27.26353804

**Authors:** Tarik Duzenli, Saliha Durmus, Hilal Esmanur Kaya, Fatih Erdoğan Sevilgen, Gulsum Kayhan, Tunahan Cakır, Mehmet Ali Ergün

## Abstract

**Background:** RNA sequencing (RNA-seq) is increasingly recognized as a complementary tool to DNA-based sequencing for improving the diagnostic yield in Mendelian disorders. However, how the diagnostic performance of RNA-seq varies across molecularly and phenotypically distinct patient subgroups remains poorly defined. This study aimed to evaluate and compare the diagnostic utility of RNA-seq across three stratified groups of patients with non-diagnostic exome sequencing.

**Methods:** We performed RNA-seq on whole blood samples from 90 patients with suspected Mendelian disease in whom clinical exome or whole-exome sequencing had failed to establish a molecular diagnosis. Patients were prospectively stratified into three groups of 30: (i) patients with a candidate variant of uncertain significance (VUS) with predicted splicing impact (Group 1), (ii) patients with a specific clinical pre-diagnosis but no identified pathogenic variant (Group 2), and (iii) patients without a specific pre-diagnosis or candidate variant (Group 3). Aberrant splicing, gene expression outliers, and allele-specific expression were analyzed using multiple bioinformatic tools and compared against a GTEx-derived control cohort.

**Results:** RNA-seq contributed to a molecular diagnosis in 29 of 88 evaluable patients (32.9%). Diagnostic yield differed substantially across groups: 82.8% (24/29) in Group 1, 6.9% (2/29) in Group 2, and 10% (3/30) in Group 3. In Group 1, RNA-seq enabled reclassification of candidate VUS through direct demonstration of aberrant splicing events. In Group 2, RNA-seq identified a somatic mosaic *ACTB* variant in a patient with Baraitser-Winter syndrome that had been missed by exome sequencing and reclassified a previously deprioritized *APPL1* VUS. In Group 3, a deep intronic pseudoexon-activating variant in *IGBP1* was identified in two siblings with severe microcephaly, providing evidence for a candidate X-linked microcephaly gene, and a pathogenic *RNU4-2* variant was detected in a patient with ReNU syndrome, a non-protein-coding gene not captured by standard exome sequencing.

**Conclusions:** RNA-seq has the highest diagnostic utility when applied to evaluate candidate splice variants identified by prior DNA testing but also provides independent diagnostic value in patients without candidate variants. The systematic comparison across stratified patient groups supports the integration of RNA-seq into clinical genomic workflows and highlights the need for standardized analytic frameworks.

## Background

DNA-based tests (whole-exome sequencing, whole-genome sequencing) have become a mainstay of clinical genetic testing for Mendelian disorders, yet their diagnostic yield remains limited. For WES, this is approximately 25–30% in large, heterogeneous rare disease cohorts and 40–60% in smaller, phenotypically homogeneous populations[1]. A major contributor to this diagnostic gap is the inability of WES to detect or functionally assess variants in non-coding regions, including deep intronic and regulatory regions, which constitute approximately 98% of the genome[2]. Whole-genome sequencing (WGS) partially addresses this limitation by capturing non-coding variants, but the interpretation of the millions of variants identified per individual remains challenging [2–4]. Critically, DNA-based methods alone do not provide direct evidence of a variant’s effect on mRNA splicing or gene expression. RNA sequencing (RNA-seq) fills this gap by enabling the direct analysis of splicing events, gene expression levels, and allele-specific expression, thereby providing functional evidence to support variant interpretation[5]. Its diagnostic utility has been demonstrated in multiple rare disease cohorts, including patients with neuromuscular[6–9] and mitochondrial disease[10, 11], as well as in more heterogeneous rare disease populations, with reported diagnostic yields ranging from 2.6% to 86.7% depending on cohort composition, tissue type, and analytic approach [12–23].

The clinical relevance of splicing analysis is underscored by the observation that an estimated 15–62% of pathogenic single-nucleotide variants causing Mendelian disease are thought to exert their effects through disruption of mRNA splicing, depending on the gene involved[24, 25]. However, splicing-associated variants located outside the canonical splice donor (+1, +2) and acceptor (-1,-2) sites are routinely deprioritized in clinical variant interpretation due to their large numbers and unknown functional consequences, despite evidence that over 80% of experimentally validated splice-disrupting variants fall outside these canonical positions[26, 27]. This discrepancy is reflected in the ClinVar database, where only 16.5% of pathogenic/likely pathogenic splicing-associated variants are annotated at non-canonical sites[28], highlighting a substantial knowledge gap. RNA-seq can directly assess the functional impact of such variants on the transcript, making it a critical tool for resolving these diagnostic uncertainties.

In this study, we applied RNA-seq to 90 patients with suspected Mendelian disease in whom clinical exome or whole-exome sequencing failed to establish a definitive molecular diagnosis. Patients were stratified into three groups of 30 based on their molecular and phenotypic characteristics: (i) patients harboring a variant of uncertain significance (VUS) with predicted splicing impact, (ii) patients with a specific clinical pre-diagnosis but no pathogenic/likely pathogenic variant in the relevant disease genes, and (iii) patients without a specific pre-diagnosis and no pathogenic/likely pathogenic variant identified by WES. This design allowed us to systematically evaluate and compare the diagnostic utility of RNA-seq across these distinct clinical scenarios — an approach that, to our knowledge, has not been previously reported.

## Methods

### Study design and cohort recruitment

This prospective cohort study enrolled 90 patients aged 2.5–59 years who were referred to the Department of Medical Genetics at Gazi University Faculty of Medicine between January 2025 and February 2026 from various clinical departments. All patients had undergone prior clinical exome or whole-exome sequencing without reaching a definitive molecular diagnosis.

Patients were stratified into three groups of 30 based on their molecular findings and clinical presentation. Group 1 comprised patients harboring a VUS predicted to affect splicing, which was also clinically concordant with the patient’s phenotype. Group 2 included patients with a specific clinical pre-diagnosis in whom clinical exome or whole-exome sequencing of the relevant disease genes failed to identify a pathogenic or likely pathogenic variant. Group 3 consisted of patients without a specific clinical pre-diagnosis in whom whole-exome sequencing did not identify any pathogenic or likely pathogenic variant (Figure 1).

**Figure 1:**
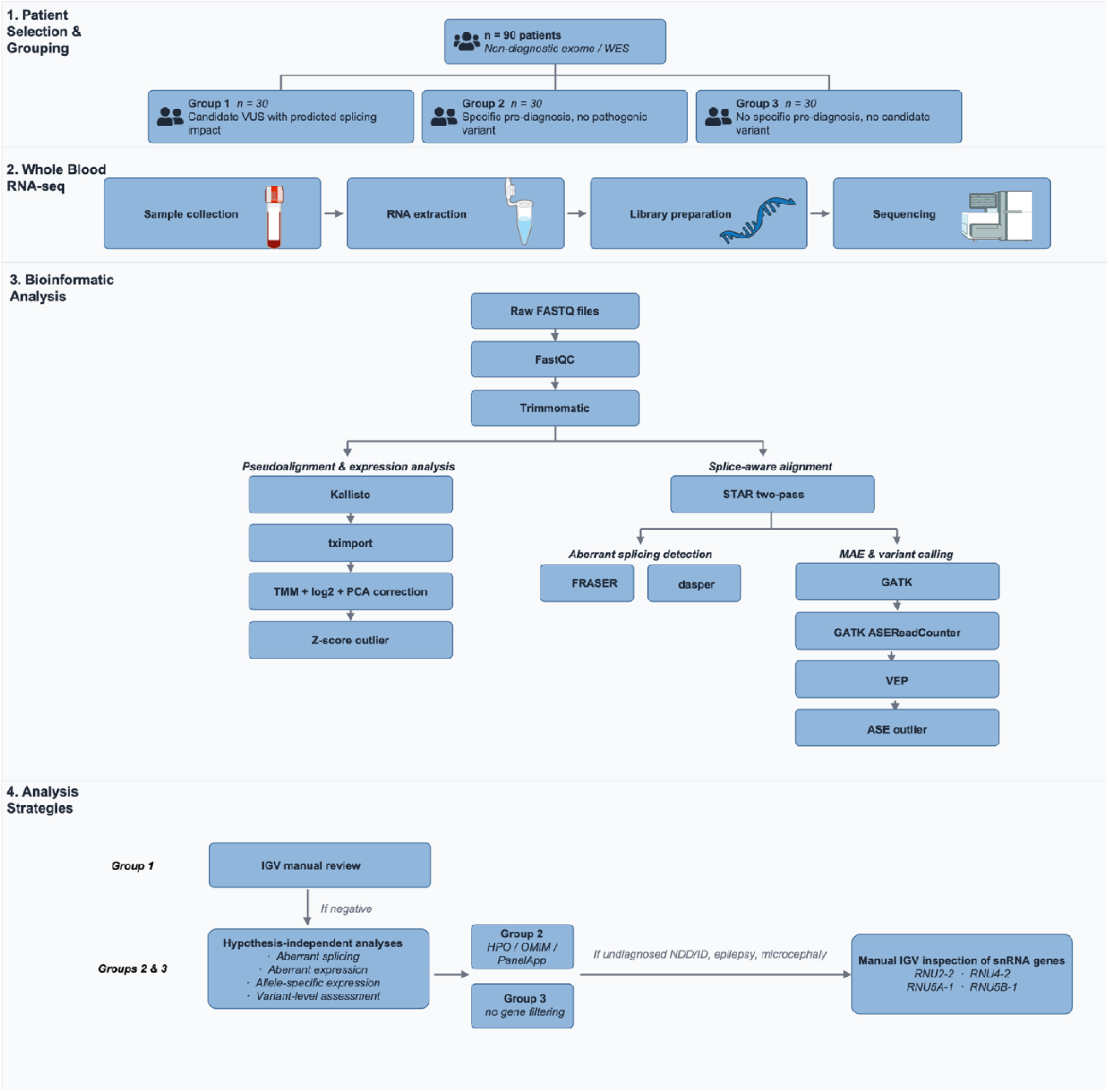
Study design and analytical workflow. Overview of the four sequential stages of the cohort study. **(1) Patient selection and grouping:** Ninety patients with suspected Mendelian disease and non-diagnostic clinical exome or whole-exome sequencing (WES) were prospectively enrolled and stratified into three groups of 30. Group 1 included patients harboring candidate variants of uncertain significance (VUS) with predicted splicing impact; Group 2 included patients with a specific clinical pre-diagnosis but no pathogenic variant identified by WES; Group 3 included patients with no specific pre-diagnosis and no candidate variant. **(2) Whole-blood RNA-seq:** Total RNA was extracted from K-EDTA whole blood, libraries were prepared with the Illumina Stranded mRNA Prep kit or NEBNext® Poly(A) mRNA Magnetic Isolation Module and paired-end sequencing was performed on the Illumina NovaSeq 6000 or MGI DNBSEQ-T7 platform **(3) Bioinformatic analysis:** Reads were quality-controlled, trimmed, and processed through two parallel branches: a pseudoalignment and expression branch (Kallisto, tximport, expression normalization, z-score outlier detection) and a splice-aware alignment branch (STAR two-pass), supporting aberrant splicing detection (FRASER, dasper), variant calling (GATK), and allele-specific expression analysis (ASEReadCounter, VEP). **(4) Analysis strategies:** Group 1 underwent targeted Integrative Genomics Viewer (IGV) review of the candidate variant locus. When negative, or for Groups 2 and 3, hypothesis-independent analyses were applied across aberrant splicing, aberrant expression, allele-specific expression, and variant-level assessment. Group 2 results were filtered against HPO/OMIM and PanelApp-derived gene panels matched to the patient’s clinical pre-diagnosis; Group 3 was analyzed without gene-level filtering. For undiagnosed patients with neurodevelopmental delay, intellectual disability, epilepsy, or microcephaly, manual IGV inspection of small nuclear RNA (snRNA) genes (RNU4-2, RNU2-1, RNU5A-1, RNU5B-1) was additionally performed. Full pipeline parameters and software versions are provided in the Methods. **Image credits:** Icons used in this figure were sourced from NIAID NIH BIOART Source (bioart.niaid.nih.gov; public domain): Blood Vial (entry 52), Eppendorf Tube (entry 143), Next Gen Sequencer (entry 386), and RNA (entry 452).

The study was approved by the Clinical Research Ethics Committee of Gazi University Faculty of Medicine (decision dated December 6, 2024; approval number 42).

### RNA extraction and library preparation

Total RNA was extracted from 10 mL of peripheral venous blood collected in potassium EDTA tubes using the QIAamp RNA Blood Mini Kit (Qiagen, Valencia, CA) according to the manufacturer’s instructions. Sequencing libraries were prepared using the Illumina Stranded mRNA Prep, Ligation Kit (Illumina Inc., San Diego, CA) or NEBNext® Poly(A) mRNA Magnetic Isolation Module (New England Biolabs, Ipswich, MA, USA) following the manufacturer’s protocol. Briefly, poly-A-tailed mRNA was purified from total RNA via poly-A capturing using oligo(dT) magnetic beads, followed by fragmentation, first-and second-strand cDNA synthesis, end repair, 3′ adenylation, adapter ligation, fragment cleanup, PCR amplification, and solid-phase reversible immobilization (SPRI) bead purification. Final library size distribution and concentration were assessed using the Agilent Bioanalyzer High Sensitivity DNA Kit (Agilent Technologies, Santa Clara, CA) and the Qubit dsDNA HS Assay Kit (Thermo Fisher Scientific, Waltham, MA), respectively. Libraries were diluted in resuspension buffer (RSB) prior to loading.

Paired-end sequencing was performed on the Illumina NovaSeq 6000 System (Illumina Inc., San Diego, CA) or MGI DNBSEQ-T7 (MGI Tech Co., Ltd., Shenzhen, China) according to the manufacturer’s instructions, targeting a minimum coverage of 50 million read pairs per sample.

### Preprocessing of raw RNAseq data

Quality control of raw sequencing data (.fastq files) was performed using FastQC (v0.11.7)[29]. Q20, Q30, and GC content metrics were calculated for each sample. Low-quality reads were trimmed using Trimmomatic (v0.39)[30], removing bases with a Phred quality score below 20 and filtering reads shorter than 50 base pairs. Post-trimming quality metrics were recalculated to ensure data integrity.

Gene-level expression quantification was performed using Kallisto (v0.46.1)[31] with the GENCODE v38 reference transcriptome (https://www.gencodegenes.org/human/release_38.html). Transcript-level abundance estimates were summarized to gene-level counts using the tximport R package (v1.22.0)[32] and pseudo-alignment rates were calculated for each sample. Trimmed reads were aligned to the GRCh38 reference genome using STAR (v2.7.10a)[33] in two-pass mode with GENCODE v38 gene annotations to enable detection of novel exon–exon junctions for downstream splicing analyses.

### RNA-seq Quality Control and Sample Exclusion

Quality control was performed using post-alignment metrics, which included a unique mapping rate ≥ 80%, exonic alignment rate ≥ 60%, and globin gene alignment rate ≤ 5%. Two samples (G2-27, G1-28) were excluded from all analyses due to failed mRNA enrichment (exonic rate 6.2%) and suspected sample contamination, respectively. A third sample (G1-6), derived from a Group 1 patient with a candidate splicing variant showed elevated globin content (33.8%) and reduced unique alignment rate (47.6%); however, as variant-supporting reads at the candidate locus were of sufficient depth and quality, this sample was retained for targeted analysis only and excluded from transcriptome-wide outlier detection and cohort-level statistics due to potential bias in count-based outlier detection. The final dataset for transcriptome-wide outlier analyses comprised 87 samples.

### Variant calling

Variant calling was performed using the Genome Analysis Toolkit (GATK, v4.6.0.0)[34]. Duplicate reads were marked with MarkDuplicates and reads containing “N” characters in the CIGAR string were split using SplitNCigarReads. Variants were called using HaplotypeCaller and filtered according to GATK-recommended hard filtering criteria. Variant annotation was performed with Ensembl VEP (v104)[35], and gene–phenotype associations were obtained from Human Phenotype Ontology (HPO)[36], ClinGen[37], PanelApp[38] and Online Mendelian Inheritance in Man (OMIM)[39].

Variants were filtered by minor allele frequency (MAF <0.01 in gnomAD v4.1.0) and prioritized by predicted functional impact, including nonsense, frameshift, canonical splice-site, stop-loss, start-loss, indel, missense, and synonymous or intronic variants predicted to affect splicing (SpliceAI Δ-score >0.2). Variants with at least one pathogenic submission in ClinVar were also retained regardless of frequency. For Group 2, prioritized variants were further filtered against the patient’s clinical phenotype using HPO-based gene panels, whereas for Group 3 no phenotype-based filtering was applied given the absence of a specific pre-diagnosis.

### Detection of aberrant events from RNA-Seq data

G&M (PhiTech, www.genomicsandmore.com), a multi-omics decision-support platform, was used to support the interpretation of genomic and transcriptomic findings. Detected aberrant splicing events, genes with aberrant expression and allele-specific expression, as described below, were visualized in G&M for interpretation. Final interpretation was performed by the clinical/genetics team.

A control dataset was assembled from the Genotype-Tissue Expression (GTEx) Consortium v8 release[40]. Whole-blood samples were selected based on the following quality criteria: RNA Integrity Number (RIN) ≥ 6, total read count ≥ 50 million, ischemic time < 720 minutes, and body mass index (BMI) < 30, yielding a final control cohort of 209 samples.

Aberrant splicing events were detected using two tools: FRASER[41] and dasper[42]. Both tools evaluate whether splice junction read counts deviate significantly from the distribution observed in the control cohort. For FRASER, statistical significance was defined as *p* < 0.05 with effect size thresholds of Δψ (delta percent spliced in) and Δθ (delta theta) > 0.15. For dasper, an isolation forest model (IFM) rank < 1,000 was used as the threshold for aberrant splicing. Aberrant splice junctions located within 500 base pairs upstream or downstream of a candidate variant were considered variant-associated.

Aberrant gene expression was defined as a significant deviation of a gene’s expression level from the reference samples. Patient expression data were merged with the control cohort and normalized using the trimmed mean of M-values (TMM) method with log2 transformation[43, 44]. Latent biological and technical covariates were identified by principal component analysis (PCA), and the principal components representing the major sources of variation were regressed out. Z-scores were calculated for each gene and transcript using the corrected expression data. A gene or transcript was classified as an expression outlier when the z-score was ≤ −2 or ≥ 2, corresponding approximately to *p* < 0.05.

Allele-specific expression (ASE) was assessed at heterozygous biallelic single-nucleotide polymorphism (SNP) positions. Allele-level read counts were quantified using GATK ASEReadCounter[34], incorporating both RNA-seq–derived and genomic variant calls. Variant annotation was performed with Ensembl VEP (v104)[35]. A gene was classified as exhibiting ASE when the mean allele balance in the patient sample fell outside the 95% confidence interval of the control cohort distribution.

### Confirmation studies

Variants of interest identified by RNA-seq were confirmed by Sanger sequencing and/or targeted next-generation sequencing (NGS) of genomic DNA. Primers flanking the target regions were designed using NCBI Primer-BLAST; primer sequences are provided in Additional file 1: Supplementary Methods (Table SM1). For Sanger sequencing, PCR products were purified using ExoSAP-IT™ PCR Product Cleanup Reagent (Applied Biosystems) and sequenced bidirectionally with the BigDye Terminator v3.1 Cycle Sequencing Kit (Applied Biosystems) on an ABI 3500 Genetic Analyzer (Applied Biosystems). Chromatograms were analyzed using SnapGene and aligned against the corresponding reference sequences. Sequencing libraries were prepared from PCR amplicons using the Illumina DNA Prep kit (Illumina, San Diego, CA, USA) according to the manufacturer’s amplicon protocol and sequenced on an MGI DNBSEQ-T7 platform. Raw reads were quality-assessed with FastQC and adapter-trimmed with Trimmomatic, and trimmed reads were aligned to the GRCh38 human reference genome using the Illumina DRAGEN Bio-IT Platform v3.9. Aligned reads were visualized in the Integrative Genomics Viewer (IGV) for variant inspection and allele fraction estimation. For patient G2-2, in whom somatic mosaicism was suspected, genomic DNA was additionally extracted from a primary fibroblast culture established from a skin punch biopsy (see Additional file 1:Supplementary Methods).

### Structural prediction of wildtype and aberrantly spliced protein isoforms

For variants predicted to result in in-frame exon skipping affecting a functionally annotated protein domain, three-dimensional structures of the wildtype and aberrantly spliced protein isoforms were predicted using ColabFold v1.5.5, [45] and visualized in PyMOL v3 (Open Source) with functional domains annotated according to UniProt. Detailed parameters and confidence assessment criteria are provided in Additional file 1:Supplementary Methods.

### Analysis strategies

For patients in Group 1, the candidate variant was first examined manually using the Integrative Genomics Viewer (IGV)[46], comparing splicing patterns to three randomly selected samples from the cohort. A minimum of 5 supporting reads was required at the region of interest for aberrant splicing event identification.

For patients in Groups 2 and 3, as well as Group 1 patients in whom the candidate variant did not disrupt splicing, hypothesis-independent analyses were performed, including aberrant splicing, aberrant expression, allele-specific expression, and variant-level assessment, as described above. In Group 2, phenotype-matched gene filters based on HPO, OMIM, and PanelApp were applied, whereas no predefined gene filtering strategy was used in Group 3. In patients with neurodevelopmental disorder/intellectual disability (NDD/ID), epilepsy, or microcephaly who remained undiagnosed after these analyses, manual inspection of IGV alignments was performed for the snRNA genes *RNU2-2* (ENST00000410396.1), *RNU4-2* (ENST00000365668.2), *RNU5A-1* (ENST00000362698.2), and *RNU5B-1* (ENST00000363286.2).

Variants were classified according to ACMG/AMP 2015[47] and ACGS 2024[48] guidelines. Aberrant splicing events were evaluated according to the recommendations of the ClinGen Sequence Variant Interpretation (SVI) Splicing Subgroup[49], and appropriate evidence criteria were applied. Identified splicing events were visualized as sashimi plots using an in-house script based on MISO[50]

## Results

### Cohort characteristics

A total of 90 patients were enrolled, aged 2.5 to 59 years (mean 17 years), of whom 53 were in the pediatric age group and 37 were adults. Forty patients (45%) were female and 50 (55%) were male. Consanguinity (within seven degrees) was present in 42 families (46.6%). The cohort included 14 sibling pairs and one father–daughter pair.

Across the entire cohort, the most frequent phenotypic category was neurological (n = 50, 55.6%), followed by multisystem involvement (n = 13, 14.4%), ophthalmological (n = 8, 8.9%), immunological (n = 5, 5.6%), dermatological (n = 4, 4.4%), and other systems including metabolic, skeletal, cardiovascular, audiological, hematological, and renal involvement at lower frequencies (Figure 2). Prior to RNA-seq, 85 patients had undergone whole-exome sequencing (WES), three had clinical exome sequencing (G1-25, G2-3, G2-15), and two had single-gene analysis (G1-29, G1-30). Clinical summaries and phenotypic details for all patients are provided in Additional file 2: Supplementary Tables S1–S3.

**Figure 2:**
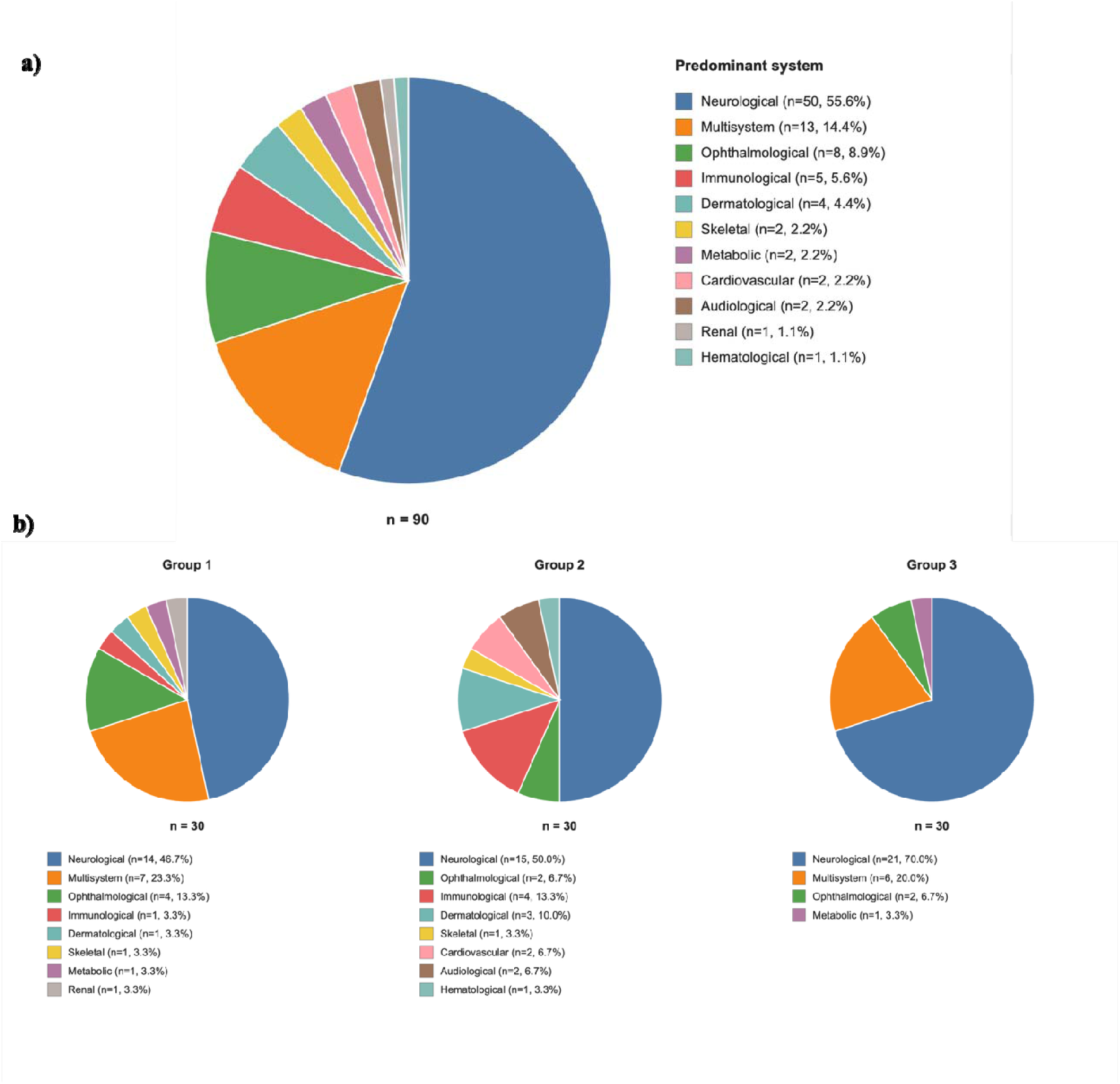
Predominant clinical system distribution across the cohort. **(a)** Pie chart of the predominant affected organ system across the entire cohort (n = 90). Neurological involvement was the largest category (n = 50, 55.6%), followed by multisystem presentations (n = 13, 14.4%), ophthalmological (n = 8, 8.9%), immunological (n = 5, 5.6%), and dermatological (n = 4, 4.4%) categories, with smaller proportions of skeletal, metabolic, cardiovascular, audiological, renal, and hematological phenotypes. **(b)** Group-specific distributions of predominant clinical systems for Group 1 (left), Group 2 (middle), and Group 3 (right; n = 30 per group). Group 3 was most strongly enriched for neurological phenotypes (n = 21, 70.0%), consistent with the inclusion of patients with unexplained neurodevelopmental disorders, intellectual disability, and epilepsy. Group 2 showed the most heterogeneous phenotypic distribution, reflecting its enrolment criterion of a specific pre-diagnosis without an identified pathogenic variant. Each patient was assigned to a single predominant system on the basis of the clinical features driving the diagnostic referral.

### Sequencing Quality Metrics

Following adapter trimming, samples (n = 87) yielded a median of 158.2 million paired-end reads (range: 75.5–678.2), with a median of 70.3 million uniquely aligned read pairs (range: 29.3–283.9) and a median unique alignment rate of 91.3% (range: 57.1–95.2%). The median exonic mapping rate was 71.5% (range: 54.5–91.9%), and the median globin transcript content was 0.13% (range: 0.00–2.52%), indicating generally efficient mRNA capture across the cohort.

### Transcriptome-wide Outlier Profiling

Transcriptome-wide outlier analyses (n = 87) identified a median of 211 splicing outlier genes per sample by FRASER (range: 0–465) and 164 by dasper (range: 0–455), with a median of 24 genes detected concordantly by both tools (range: 0–61). Expression outlier analysis identified a median of 1,518 outlier genes per sample (range: 4–4,378), and monoallelic expression (MAE) analysis identified a median of 153 genes per sample (range: 0–836). A median of 11,652 genes per sample were expressed at TPM ≥ 1 (range: 7,014–12,100), establishing the analytical coverage available for clinical interpretation.

### Group 1: Hypothesis-driven splicing analysis of candidate variants of uncertain significance

Of the 30 patients in Group 1, one (G1-28) was excluded due to suspected sample contamination. Among the remaining 29 patients, RNA-seq demonstrated that the candidate VUS disrupted splicing in 24 cases (82.8%), leading to reclassification as pathogenic or likely pathogenic. In four patients (G1-4, G1-12, G1-17 and G1-23) no effect on splicing was observed, and in one patient (G1-3), insufficient read coverage at the region of interest precluded assessment. The candidate genes had median TPM values in GTEx ranging from 0.21 to 182.7, and SpliceAI scores ranged from 0.01 to 0.99 (Table S1.)

Among the 24 positive cases, the observed aberrant splicing events included exon skipping (n=9), alternative acceptor site usage (n=3), alternative donor site usage (n=3), intron retention (n=2), and combined events (n=7). SpliceAI correctly predicted the type of splicing disruption with full accuracy in 54.1% of cases (13/24), partial accuracy in 29.1% (7/24), and made an incorrect prediction in 16.7% (4/24). On the other hand, among aberrant splicing detection tools, dasper identified 15 while FRASER detected 5 of the 24 positive events. In one case (G1-29), FRASER detected a meaningful effect size (Δψ:-0.18), but it did not reach statistical significance (p = 0.064).

Among the 19 patients harboring variants predicted to trigger NMD, a marked reduction in total transcript abundance (z-score <-2) was observed in 4/19 (21.1%), all of whom carried the variant in a bi-allelic state (3 homozygous, 1 hemizygous); no heterozygous carrier of an NMD-predicted variant showed comparable depletion. Conversely, one heterozygous patient (G1-10) carrying a variant not predicted to elicit NMD nonetheless showed reduced expression (z =-2.34).

Complete results for all patients with aberrant splicing events across Groups 1–3 are summarized in Table 1 and Additional file 3: Supplementary Figures S1–S22. Below, we highlight selected cases illustrating distinct molecular mechanisms elucidated by RNA-seq in this group.

**Table 1.**
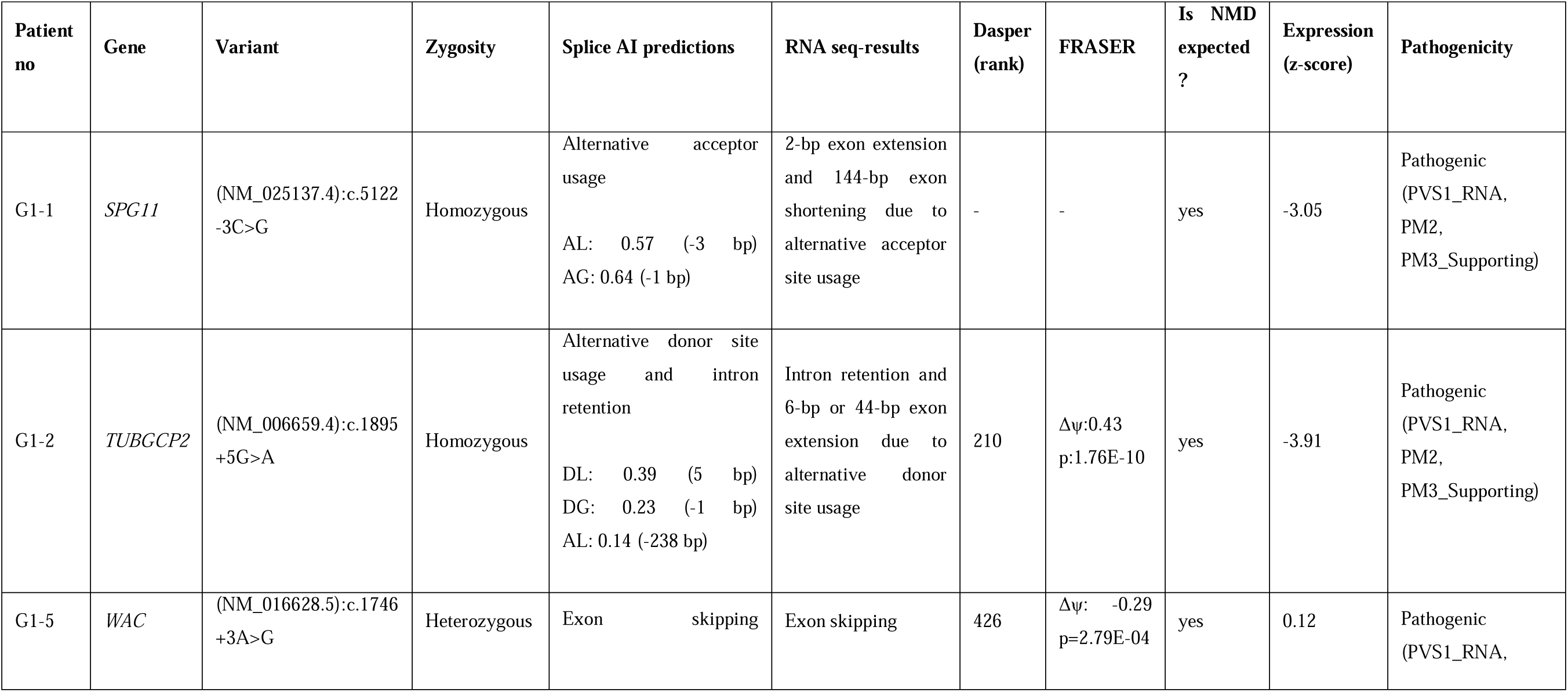
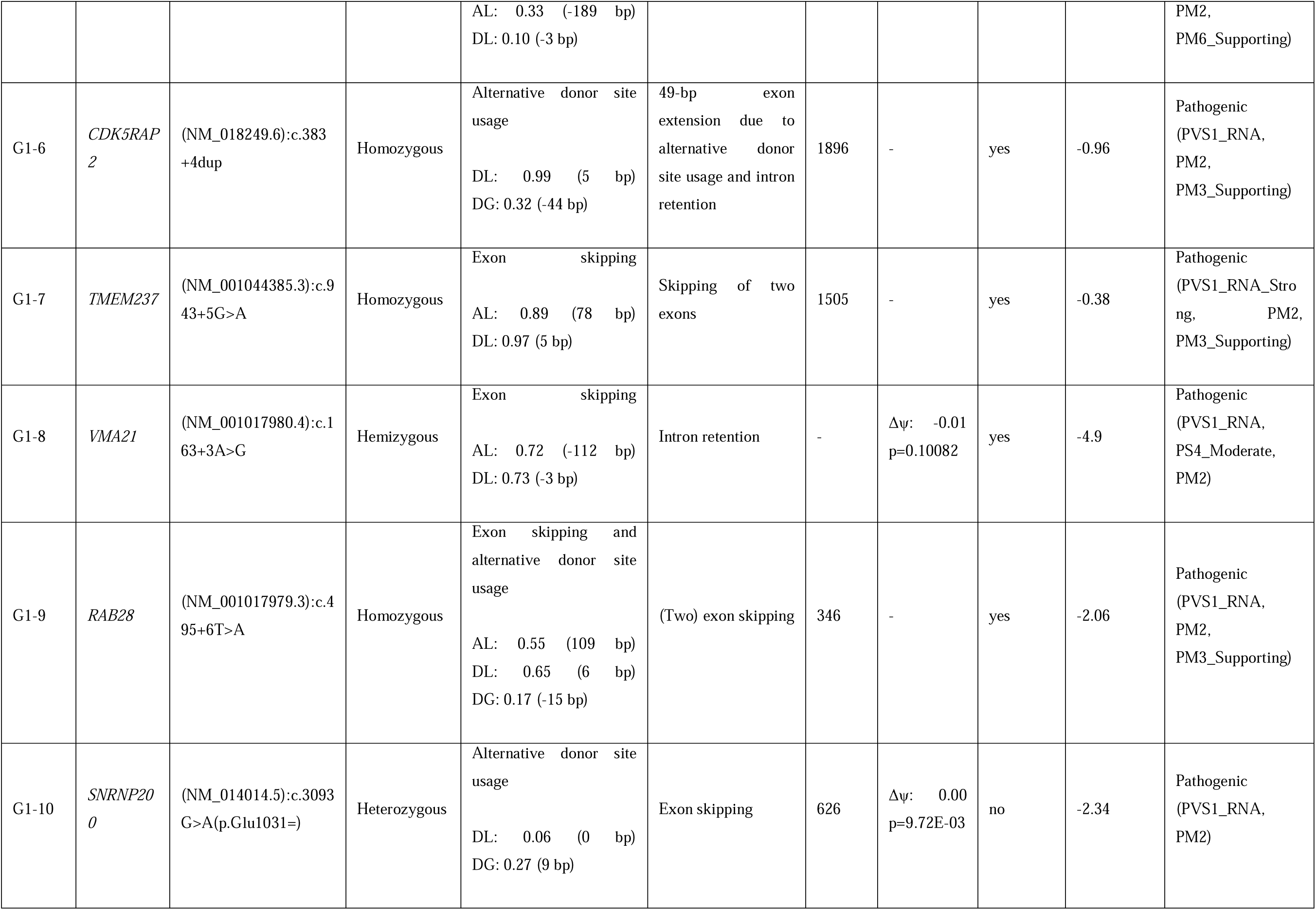
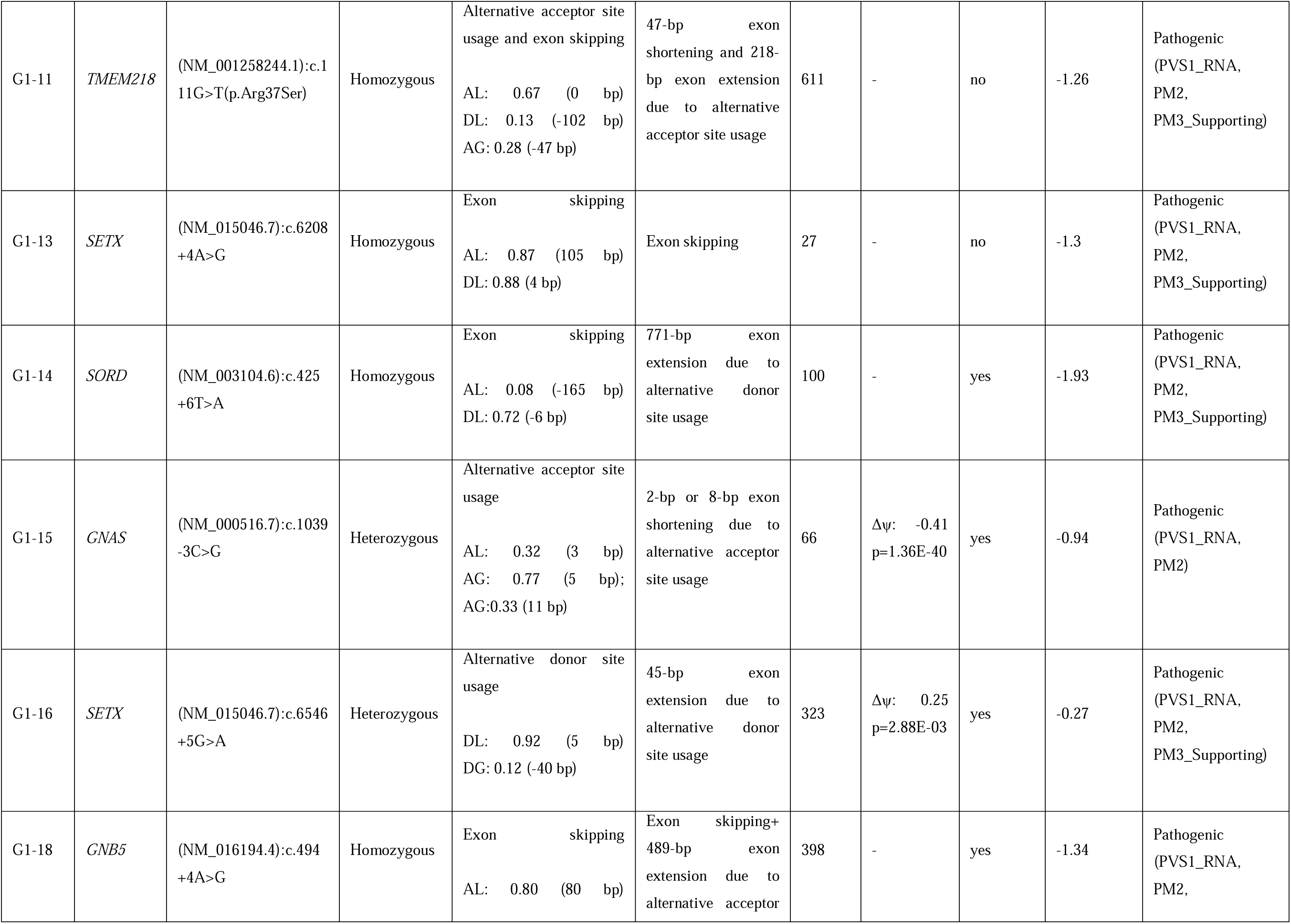
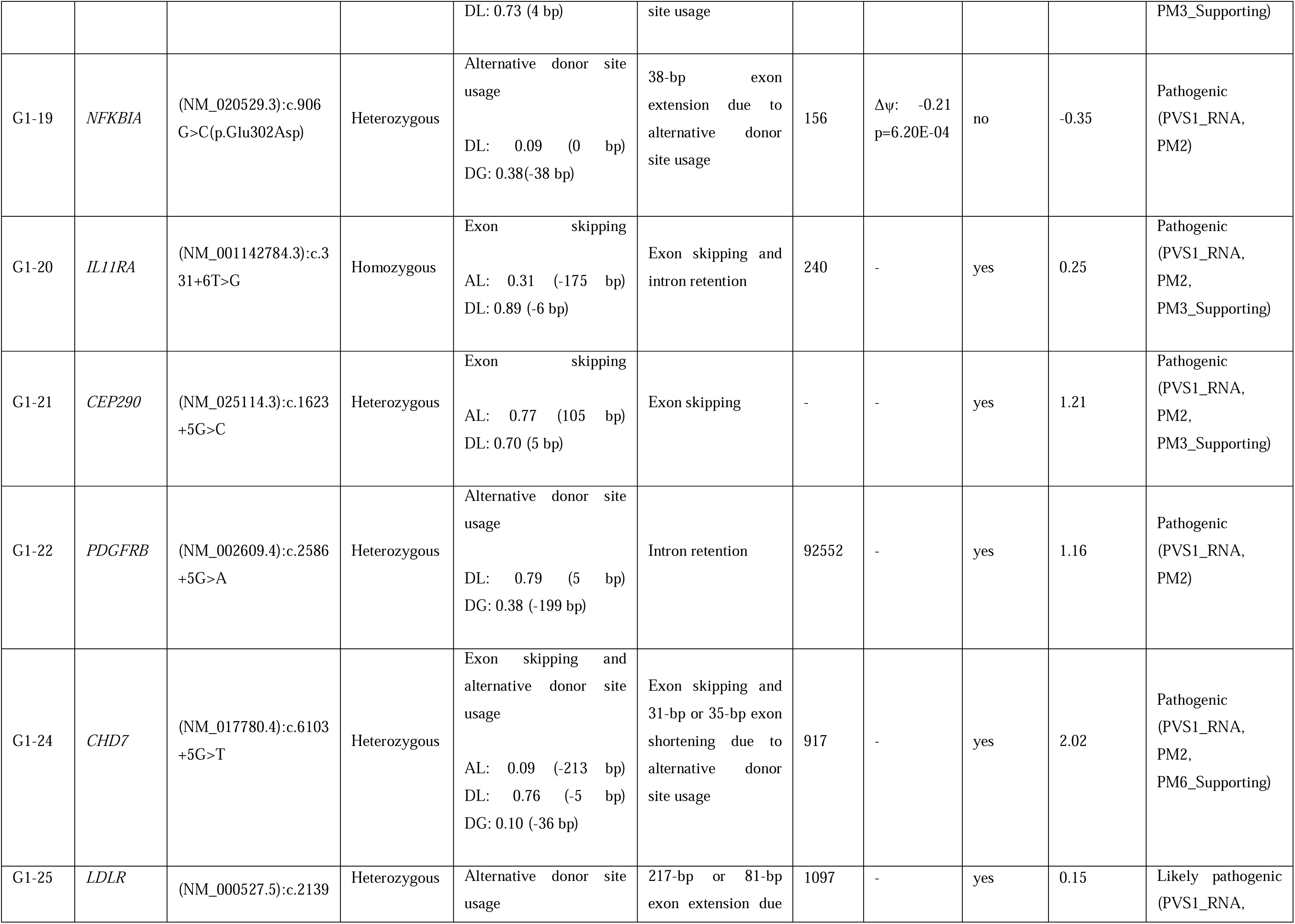
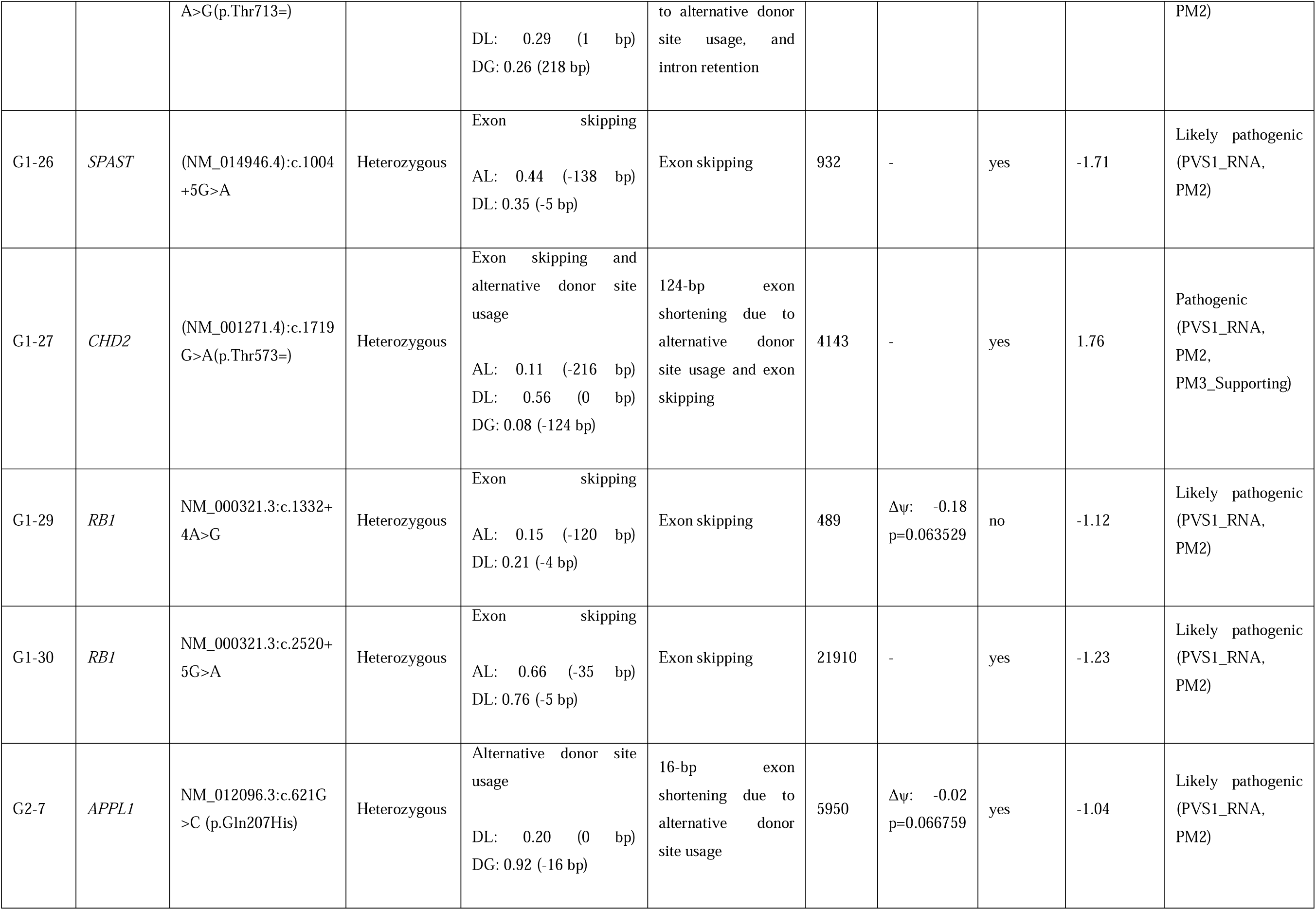
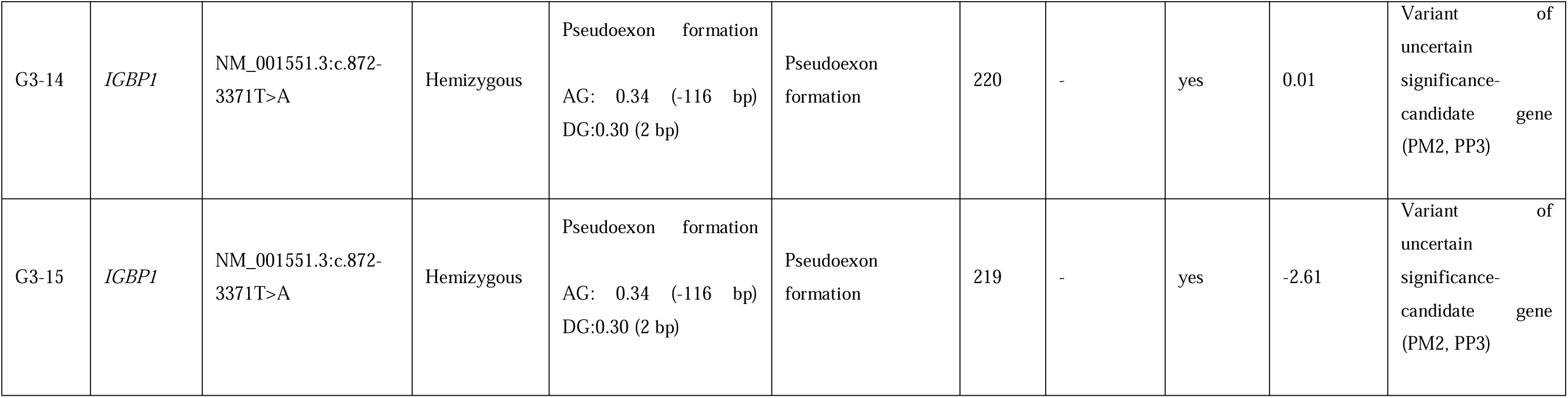
RNA-seq–confirmed aberrant splicing events in 27 patients reaching a molecular diagnosis through demonstration of splicing aberration. For each patient, the affected gene, candidate variant (HGVS nomenclature), zygosity, SpliceAI predictions, observed RNA-seq splicing event, results of transcriptome-wide aberrant splicing detection (dasper isolation forest model rank and FRASER Δψ with associated p-value), predicted susceptibility to nonsense-mediated decay (NMD), gene-level expression z-score, and final variant classification with applied evidence criteria are reported. SpliceAI Δ-scores are shown for acceptor loss (AL), acceptor gain (AG), donor loss (DL), and donor gain (DG), with the predicted distance to the canonical site indicated in parentheses. A dash (–) indicates that no event was detected at the variant locus by the corresponding tool.

#### G1-1

G1-1 was a male patient in the 31–35 age range who presented with progressive gait disturbance beginning in the third decade, numbness in the upper extremities, dysarthria, and macular retinal dystrophy. Family history revealed parental consanguinity and an affected sibling with onset of gait disturbance in the third decade. Physical examination showed bilateral lower-extremity spasticity, hyperactive deep tendon reflexes, clonus, and bilateral paraparesis (3/5). The patient demonstrated a broad-based, spastic gait pattern. Electromyography was normal. Visual evoked potential testing showed reduced amplitudes consistent with bilateral dysfunction. Cranial MRI revealed focal hyperintense signal changes on T2-and FLAIR-weighted images in the bilateral centrum semiovale and periventricular white matter, together with a thin corpus callosum.

Exome sequencing identified a homozygous c.5122-3C>G variant in the *SPG11* gene (NM_025137.4). This variant had not been reported in population databases of healthy individuals (gnomAD v4.1.0) or in the literature. Based on the available evidence, it was classified as a variant of uncertain significance (PM2, PP3, PM3). SpliceAI predicted a two-base exon extension due to alternative acceptor site usage (acceptor loss [AL]: 0.57 [-3 bp]; acceptor gain [AG]: 0.64 [-1 bp]).

Inspection of RNA sequencing data in IGV demonstrated that the variant disrupted the canonical acceptor site, while the variant position together with the immediately upstream nucleotide functioned as a novel acceptor site. This alteration resulted in a two-base (CT) extension of exon 30, which appeared in IGV as an insertion within exon 29 because the last four nucleotides of exon 29 also end with CT (Figure 3a). Thus, this variant was interpreted as a duplication event and could not be detected by aberrant splicing tools. In addition, a small fraction of reads demonstrated another event absent in control samples, in which a 144-bp deletion in exon 30 was identified due to the use of an alternative acceptor site (Figure 3b).

**Figure 3:**
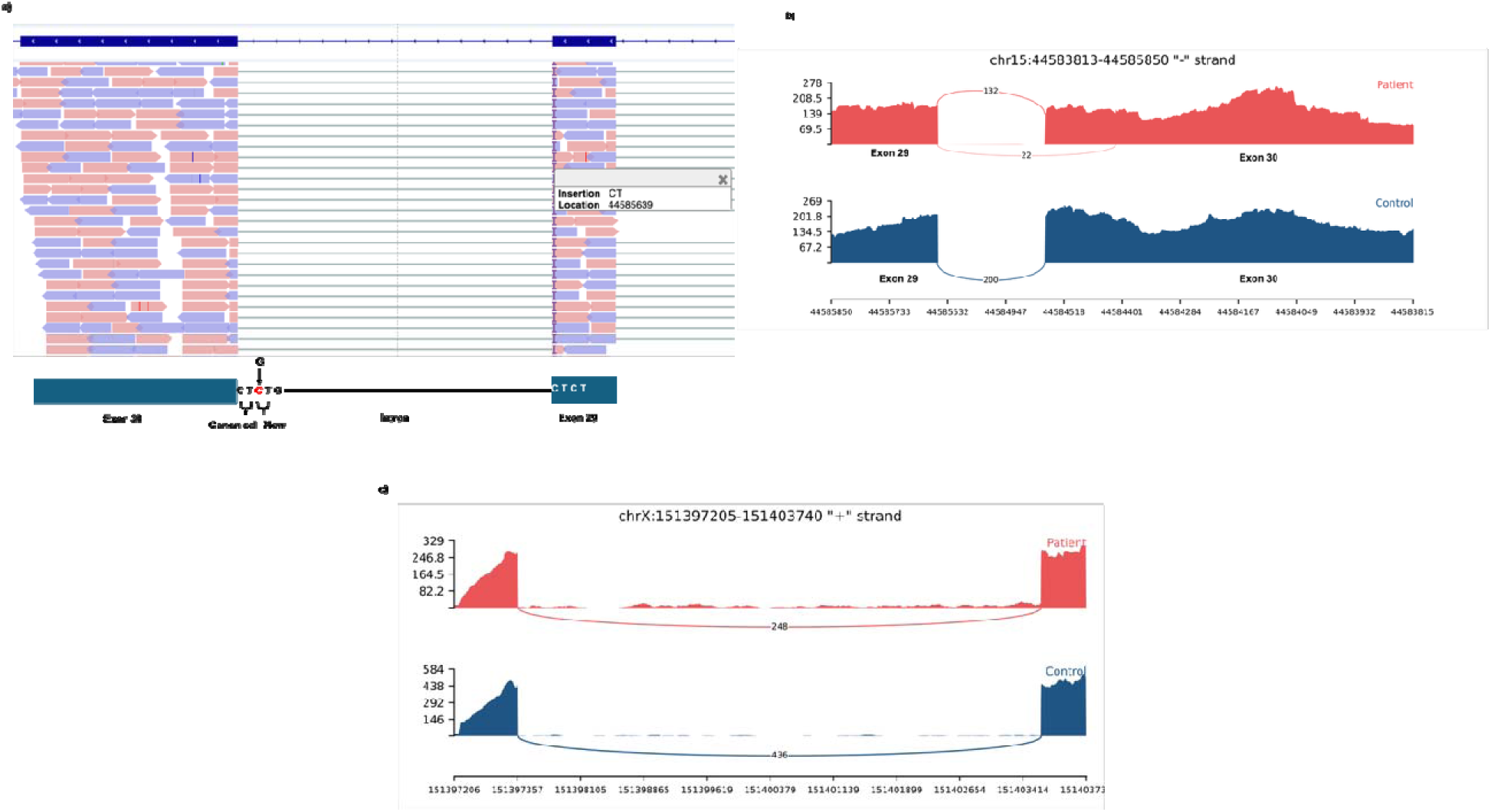
RNA-seq confirmation of splice-impacting candidate variants in three Group 1 patients. **(a)** Integrative Genomics Viewer (IGV) view of the *SPG11* locus in patient G1-1 (homozygous c.5122-3C>G). Reads support the use of an alternative acceptor site, with the original canonical acceptor and a novel acceptor positioned upstream as schematized below the read alignment, producing a 2-bp exon extension (r.5121_5122insAG) and a 144-bp exon shortening (r.5122_5266del). **(b)** Sashimi plot of the same locus in the patient (red) compared with a control sample (blue), showing the alternative junction (22 reads, patient) absent from the control (200 reads on the canonical junction). **(c)** Sashimi plot of the *VMA21* locus (chrX:151,397,205–151,403,740) in patient G1-8 (red) versus a control (blue). The patient shows minimal intron retention, leading to r.163_164ins163+1_164-1, p.(Ala56Asnfs*14). A pedigree of the family of patient G1-8 has been omitted to protect family privacy and is available from the corresponding author on reasonable request.

The variant was predicted to disrupt the reading frame generate a premature termination codon (r.[5121_5122insAG], [5122_5266del]; p.[(Ile1708Argfs*2)], [(Ile1708_Ala1756del)]), thereby triggering nonsense-mediated decay (NMD). Consistent with this, expression analysis showed a z-score of −3.05 for *SPG11* gene, indicating markedly reduced gene expression. According to current guidelines, the variant was reclassified as pathogenic based on the criteria PVS1_RNA, PM2, and PM3_Supporting. This established the molecular diagnosis in the patient.

#### G1-8

A male patient in the 26–30 age range was referred to our clinic because of easy fatigability, gait disturbance, and muscle weakness with onset in the second decade of life. His parents were non-consanguineous. Similar complaints were reported in multiple relatives, and the proband’s female child was noted to have lower-extremity muscle pain and gait disturbance. Muscle biopsy from proband demonstrated myopathic/dystrophic changes, while electromyography revealed widespread myogenic involvement consistent with myopathy. Based on the clinical findings, calpainopathy/limb-girdle muscular dystrophy was initially suspected.

Exome sequencing identified a hemizygous c.163+3A>G variant in the *VMA21* (NM_001017980.4). This variant has not been reported in population databases (gnomAD v4.1.0). It was previously described in a family suspected of having X-linked myopathy[51] and based on the available evidence, had been classified as a variant of uncertain significance (PM2, PP3, PS4_Moderate). The same variant was also detected in the proband’s female child, whereas testing has not yet been performed in the other affected relative. SpliceAI predicted skipping of exon 2 (AL: 0.72 [-112 bp]; donor loss [DL]: 0.73 [-3 bp]).

RNA sequencing analysis demonstrated partial retention of intron 2 in a subset of reads (Figure 3c). This event was predicted to disrupt the reading frame and generate a premature termination codon (r.163_164ins163+1_164-1, p.(Ala56Asnfs*14)), thereby triggering NMD. Expression analysis further supported this finding, showing a *VMA21* z-score of −4.9. Complete loss-of-function variants in *VMA21* are considered incompatible with life, whereas hypomorphic variants are known to cause disease[52]. The relatively low proportion of intron retention observed in our patient was therefore considered compatible with a hypomorphic effect. This event was also not detected by both aberran splicing tools.

According to current guidelines, this variant was reclassified as pathogenic based on the PVS1_RNA, PS4_Moderate, and PM2 criteria, thereby establishing the patient’s molecular diagnosis. Although clinical manifestations have not previously been reported in female carriers, the mild symptoms observed in the proband’s female child were considered potentially related to this X-linked variant, possibly as a consequence of skewed X-chromosome inactivation.

#### G1-29

A male patient in the 6–10 age range was referred to our clinic following a diagnosis of unilateral, unifocal retinoblastoma in infancy. There was no parental consanguinity, and no family history of retinoblastoma was reported. Sequencing of the *RB1* gene (NM_000321.3) identified a heterozygous c.1332+4A>G variant. This variant has not been reported in gnomAD (v4.1.0) and has been submitted by a single laboratory to ClinVar as a variant of uncertain significance. Segregation analysis demonstrated that the variant was inherited from the clinically unaffected parent (Figure 4a). Based on the available evidence, the variant was initially classified as a variant of uncertain significance (PM2, PP3). SpliceAI predicted skipping of exon 13 with modest scores (AL: 0.15 [-120 bp]; DL: 0.21 [-4 bp]).

**Figure 4:**
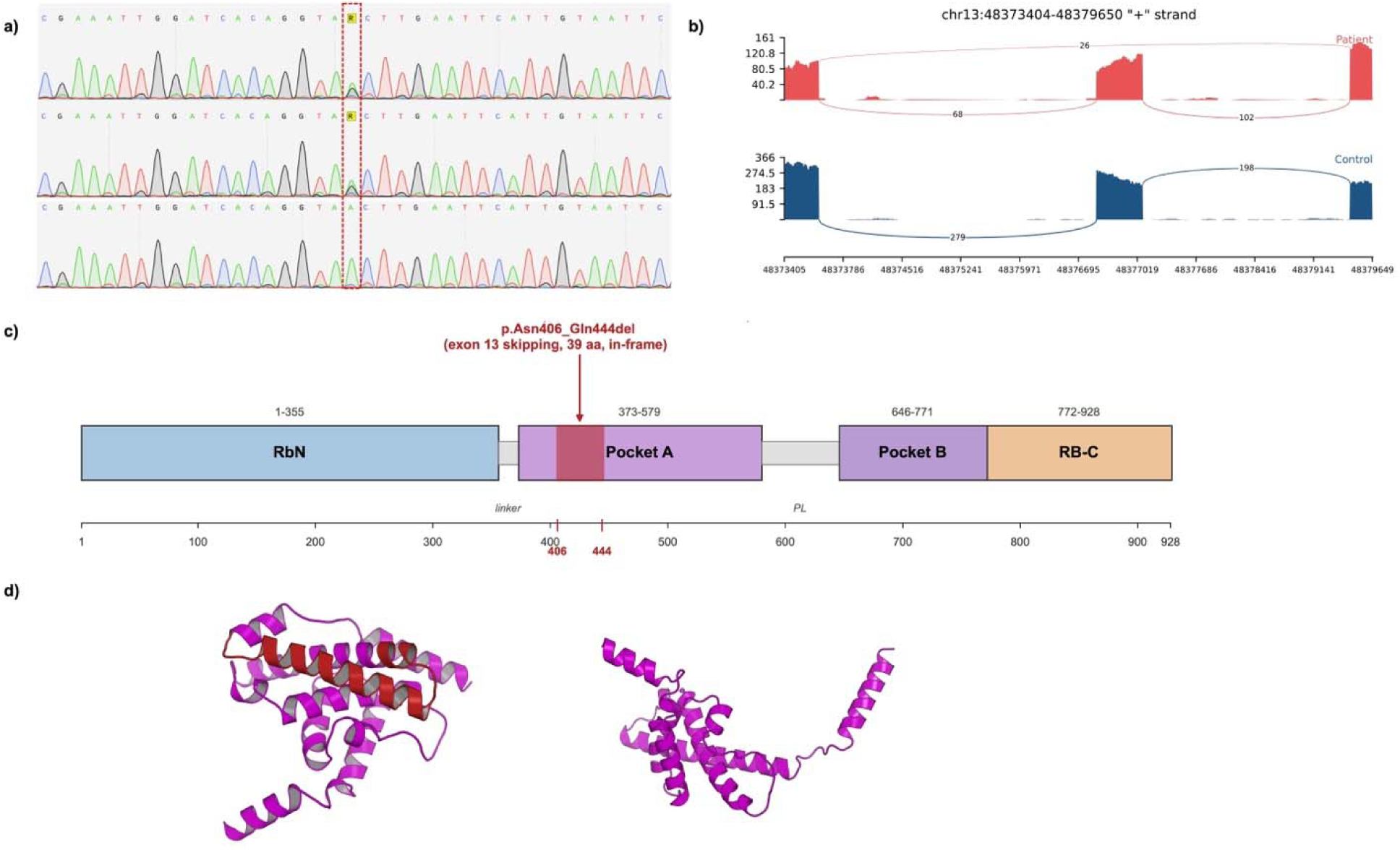
Multi-modal characterization of an *RB1* splice variant in a patient with unilateral retinoblastoma (G1-29). **(a)** Sanger sequencing chromatograms of genomic DNA from the patient (top), the clinically unaffected father (middle), and the mother (bottom), confirming paternal inheritance of the heterozygous c.1332+4A>G substitution at the position highlighted in the dashed box. **(b)** Sashimi plot of the *RB1* locus (chr13:48,373,404–48,379,650) for patient G1-29 (heterozygous NM_000321.3:c.1332+4A>G; red) compared with a control sample (blue). The patient shows a junction supporting exon 13 skipping absent from the control track. **(c)** Schematic of the RB1 protein (UniProt P06400, 928 amino acids) showing its domain architecture: RbN (residues 1–355), Pocket A (373–579), Pocket B (646–771) and the C-terminal region RB-C (772–928). The in-frame deletion p.(Asn406_Gln444del) (39 amino acids, exon 13 skipping) is mapped in red onto Pocket A. **(d)** Predicted three-dimensional structures of the wildtype Pocket A domain (left) and the in-frame deletion product p.(Asn406_Gln444del) (right), generated using ColabFold and rendered in PyMOL. In the wildtype model, the 39 deleted residues are highlighted in red and form three α-helices located within the structural core of the domain. In the mutant model, the central α-helical core is preserved; however, two flanking α-helices that are tightly packed within the canonical Pocket A fold (predicted with high confidence; pLDDT > 90) become displaced from the bundle, indicating disruption of the native fold rather than simple shortening of the protein.

Analysis of the RNA-seq data confirmed skipping of exon 13 (r.1216_1332del; p.(Asn406_Gln444del)); however, the aberrant transcript accounted for only approximately 28% of reads spanning this junction, indicating leaky splicing with substantial preservation of the canonical transcript (Figure 4b). Exon 13 comprises 117 nucleotides, and its skipping maintains the reading frame; consequently, the resulting transcript is not predicted to be targeted by NMD. Consistent with this, the expression z-score for *RB1* was only mildly reduced (−1.12). The 39 deleted residues nonetheless lie entirely within the RB-A pocket domain, a critical functional region whose integrity is required for binding of E2F transcription factors and for tumour-suppressor activity (Figure 4c)[53, 54].

To assess the structural impact of the in-frame deletion, predicted three-dimensional structures of the wildtype and aberrantly spliced RB1 isoforms were generated using ColabFold. The wildtype RB-A pocket adopted the expected compact α-helical bundle, with the 39 deleted residues mapping to three core α-helices located within the structural interior of the domain (Figure 4d, left). In the in-frame deletion model, the central α-helical core of RB-A was preserved; however, two α-helices flanking the deletion junction (predicted with high confidence; pLDDT > 90) became displaced from the canonical fold (Figure 4d, right), indicating that loss of the 39 internal residues disrupts the structural integrity of the pocket rather than producing a simple shortening of the protein.

The combination of an in-frame, NMD-escaping transcript and partial retention of canonical splicing — together producing a hypomorphic rather than complete loss-of-function allele — provides a plausible molecular explanation for the incomplete penetrance observed in this family. Such low-penetrance RB1 alleles are well recognized and have been reported to cause unilateral and unifocal retinoblastoma, as observed in our patient, in contrast to the bilateral, multifocal disease typically associated with classical null alleles[55, 56].

Based on current guidelines, the variant was reclassified as likely pathogenic (PVS1_RNA, PM2). Full PVS1 strength was retained on the basis that the in-frame deletion removes 39 residues forming an integral part of the RB-A pocket helical bundle and is predicted to disrupt the native fold of this critical functional domain.

#### Group 2: RNA-seq in patients with a specific clinical pre-diagnosis but no identified pathogenic variant

Among the 30 patients in Group 2, one was excluded due to insufficient sequencing quality. RNA-seq contributed to a molecular diagnosis in two patients (6.9% of those analyzed). Notably, the two diagnoses illustrated qualitatively different roles for RNA-seq in this group: in one patient (G2-2), RNA-seq supported the clinical pre-diagnosis through detection of a low-level mosaic variant in the expected gene, whereas in the other (G2-7), RNA-seq did not support the referring pre-diagnosis but reclassified a previously deprioritized VUS in an unrelated disease gene that accounted for part of the clinical picture. Neurological phenotypes were the most common in this group (50%, n = 15), followed by immunological (13%, n = 4) and dermatological presentations (10%, n = 3).

#### Case G2-2 — somatic mosaicism in *ACTB*

A male child in the 0–5 age range presented with dysmorphic facial features, mild developmental delay, and vesicoureteral reflux. Craniofacial examination revealed a prominent metopic suture, coarse facial features, hypertelorism, downslanting palpebral fissures, proptosis, ptosis, a broad nasal root, depressed nasal bridge, long philtrum, drooling and narrow palate, consistent with Baraitser–Winter syndrome. Cranial MRI, EEG, and echocardiography were unremarkable. Previous karyotyping, array CGH, and WES had not identified a pathogenic variant.

RNA-seq did not reveal splicing or expression abnormalities in *ACTB* or *ACTG1*; however, variant calling from the RNA-seq data identified the well-established pathogenic variant *ACTB* (NM_001101.5):c.359C>T p.(Thr120Ile) at an allele fraction of approximately 11% across 96,899 reads (Figure 5a) [57–59]. Retrospective review of the WES data confirmed the same variant at a 7% allele fraction, below the threshold for variant calling.

**Figure 5:**
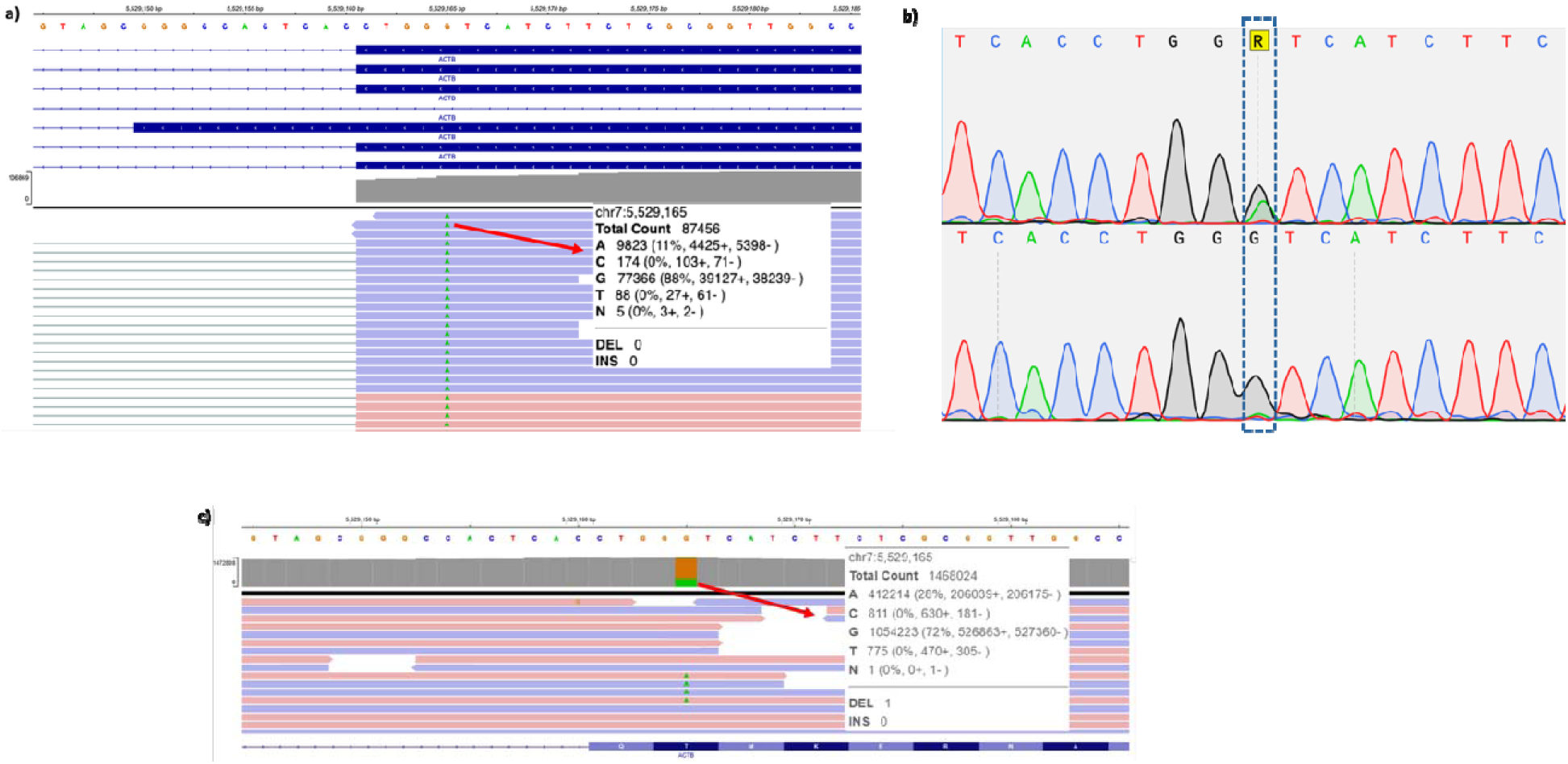
Detection of a somatic mosaic *ACTB* variant in a Group 2 patient with Baraitser–Winter syndrome. **(a)** RNA-seq IGV view of the *ACTB* locus at chr7:5,529,165 in whole-blood RNA. The base composition pile-up reveals an A allele at 11% (9,823/87,456 reads) against a reference G allele at 88%, supporting a low-level mosaic missense substitution that had not been detected on the patient’s previous clinical exome sequencing. **(b)** Sanger sequencing chromatograms of the patient at the same genomic position, comparing skin-derived DNA (top), in which the variant peak is markedly more prominent, with blood-derived DNA (bottom), in which a low-level G>A substitution is barely detectable. **(c)** Targeted next-generation sequencing of skin-derived DNA at the same position shows the variant at 28% allelic fraction (412,214/1,468,024 reads). Clinical photographs of the patient have been omitted to protect patient privacy and are available from the corresponding author on reasonable request.

Sanger sequencing of blood-derived DNA showed only a faint alternative peak at this position, whereas the variant signal was much more prominent in DNA obtained from fibroblast culture (Figure 5b). Targeted deep NGS performed on fibroblast-derived DNA identified approximately 206,000 variant reads out of 1.4 million total reads (variant allele fraction ∼28%). (Figure 5c). These findings are consistent with somatic mosaicism and may explain the patient’s relatively mild phenotype compared with typical germline Baraitser–Winter syndrome cases, despite this variant having previously been associated with a more severe clinical presentation[60]. While somatic mosaic ACTB variants have previously been associated with conditions such as Becker nevus and congenital smooth muscle hamartoma with or without hemihypertrophy, to our knowledge, somatic mosaicism manifesting as a mild Baraitser–Winter syndrome phenotype has not been reported to date.

#### Case G2-7 — RNA-seq–driven reclassification of an *APPL1* VUS in a patient referred with a CARASIL pre-diagnosis

A male patient in the 41–45 age range, born to consanguineous parents, was referred with a clinical pre-diagnosis of CARASIL. He presented with long-standing white matter disease and atherosclerotic irregularities on cranial MR angiography, sudden complete alopecia, distal paresthesias, hypothyroidism, scoliosis, type 2 diabetes mellitus, and a myocardial infarction one year before presentation; a male relative had also suffered a myocardial infarction in the fifth decade. The combination of early-onset white matter disease and alopecia supported CARASIL/CADASIL as the leading hypothesis, while the unusually early multi-system involvement broadened the differential to include progeroid syndromes.

WES did not identify a pathogenic or likely pathogenic variant in genes associated with the clinical pre-diagnosis, and RNA-seq similarly revealed no splicing or expression abnormalities in pre-diagnosis–related genes. However, an unbiased re-evaluation of remaining VUS prioritized a previously deprioritized variant in *APPL1* (NM_012096.3):c.621G>C p.(Gln207His), a gene associated with MODY type 14, on the basis of high SpliceAI scores predicting alternative donor site usage (DL: 0.20 [0 bp]; DG: 0.92 [-16 bp]).

Inspection of the RNA-seq data in IGV revealed that approximately 15% of reads at this junction utilized the alternative donor site, resulting in a 16-bp truncation of exon 8 and a frameshift introducing a premature termination codon (r.606_621del; p.(Gly202Glufs*2)) (Additional file 3: Supplementary Figure S22). This event was not detected by either FRASER or dasper. The expression z-score for *APPL1* was −1.04. The detection of the aberrant splicing event in approximately 15% of total reads is thought to reflect partial elimination of this isoform through NMD due to the premature termination codon. Since no NMD inhibitor was used in this study, the observed proportion likely underestimates the true level of aberrant transcript production.

Importantly, the molecular finding in *APPL1* did not correspond to the original CARASIL pre-diagnosis and accounted for only part of the patient’s phenotype. Upon re-evaluation of the patient’s clinical presentation, the atherosclerotic irregularities observed on cranial MR angiography, the history of myocardial infarction, and the extremity paresthesias may be attributable to poorly controlled diabetes. However, given that hyperglycemia has been present for only the past two years, whereas the vascular and neurological findings have persisted for a substantially longer period, this explanation alone does not appear sufficient. This case illustrates that in patients with a specific clinical pre-diagnosis, RNA-seq may yield molecular findings that revise rather than confirm the referring hypothesis, and that such findings may explain only a subset of the clinical features — an outcome that should be transparently reported to the patient and considered in pre-test counselling.

#### Group 3: RNA-seq in patients without a specific pre-diagnosis or candidate variant

Among the 30 patients in Group 3, RNA-seq analysis led to a molecular diagnosis in three patients (10%). Two of these were siblings (G3-14 and G3-15) sharing the same genetic etiology. Neurological phenotypes predominated in this group (70%, n = 21), followed by multisystem involvement (n = 6).

#### Cases G3-14 and G3-15 — identification of a deep intronic pseudoexon in *IGBP1*

Two affected siblings, one in the 11–15 age range and the other in the 6–10 age range, presented with severe congenital microcephaly (head circumference −7 and −7.4 SDS, respectively), global developmental delay, and recurrent infections. Both were detected prenatally in the third trimester. There was no parental consanguinity, but an affected male relative on the maternal side had similar clinical features, suggesting X-linked inheritance. Cranial MRI in the older sibling showed corpus callosum agenesis, bilateral periventricular and subcortical white matter T2 hyperintensities, and bilateral optic nerve hypoplasia. Previous microarray and WES were non-diagnostic in both siblings.

RNA-seq identified a hemizygous deep intronic variant in *IGBP1* (NM_001551.3): c.872-3371T>A, associated with the inclusion of a 119-base pair pseudoexon (r.871_872ins872-3487_872-3369; p.(Glu291Glyfs*18)). While minimal pseudoexon inclusion was observed in control samples, approximately half of the reads spanning exon 6 in both siblings were redirected to the pseudoexon (Figure 6a). The non-multiple-of-three insertion is predicted to cause a frameshift and premature termination codon, with the resulting transcript expected to undergo nonsense-mediated decay. The expression z-score was −2.61 in the older sibling and 0.01 in the younger sibling. Dasper detected this event in both siblings (IFM rank 220 and 219, respectively). The variant was confirmed by Sanger sequencing and is absent from gnomAD.

**Figure 6:**
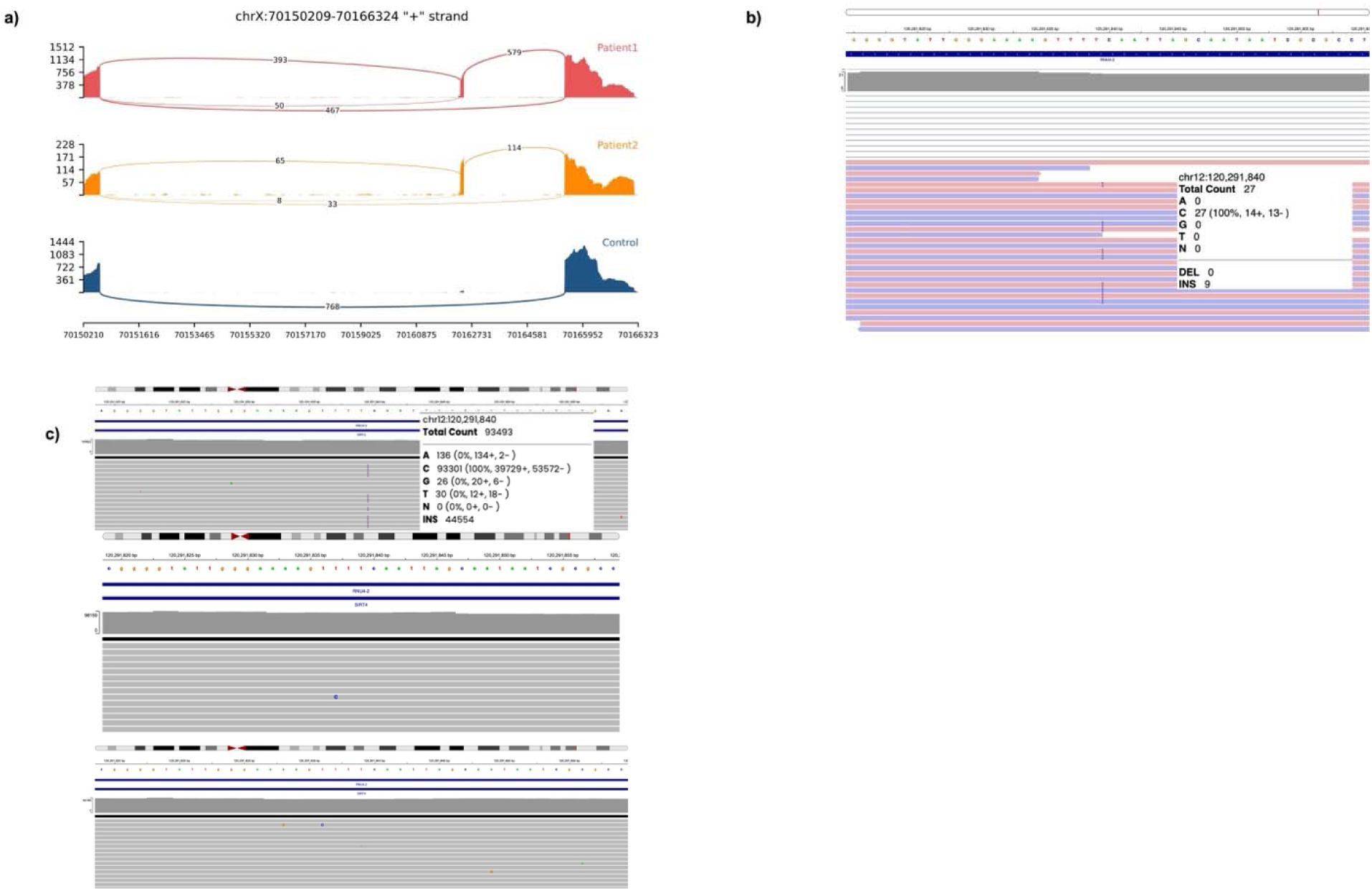
Group 3 diagnoses identified by hypothesis-independent transcriptomic analysis. **(a)** Sashimi plot of the *IGBP1* locus (chrX:70,150,209–70,166,324) in two affected brothers (G3-14, Patient 1, red; G3-15, Patient 2, orange) and a control (blue). Both patients carry the hemizygous deep-intronic variant NM_001551.3:c.872-3371T>A and show a 119-bp pseudoexon insertion supported by junction reads bridging the cryptic exon that is entirely absent from the control track (768 reads on the canonical junction). The pseudoexon predicts r.871_872ins872-3487_872-3369, p.(Glu291Glyfs*18). **(b)** RNA-seq IGV view of the *RNU4-2* locus (chr12:120,291,840) showing the n.64_65insT variant. **(c)** Targeted next-generation sequencing IGV views at the *RNU4-2* locus (chr12:120,291,840) in the proband (top) and both parents (middle and bottom), confirming the n.64_65insT variant as de novo.

*IGBP1* is listed in OMIM in association with corpus callosum agenesis with impaired intellectual development, ocular coloboma, and micrognathia; however, this association is based on a single historical report [61] and this gene-disease association was classified as “disputed” by the ClinGen Intellectual Disability and Autism Gene Curation Expert Panel. No hemizygous null variants have been reported in gnomAD and the LOEUF score is 0.24.

*IGBP1* (also known as α4) encodes a regulatory subunit that binds and stabilizes the catalytic subunits of protein phosphatase 2A (PP2A), PP4, and PP6 by protecting them from polyubiquitination until they are incorporated into functional holoenzymes[62, 63]. Notably, *de novo* pathogenic variants in other members of this phosphatase family — *PPP2R5D*, *PPP2R1A*, *PPP2CA*, and more recently *PPP2R5C* — cause the Houge–Janssens syndrome spectrum (HJS1–4), a group of well-established Mendelian neurodevelopmental disorders characterized by intellectual disability, hypotonia, macrocephaly and microcephaly in some cases[64], supporting the biological plausibility of IGBP1 as a disease gene within the same pathway. Based on the current clinical, molecular, and segregation evidence, *IGBP1* is a strong candidate gene for X-linked primary microcephaly, and functional validation studies using *C. elegans* models have been initiated.

#### Case G3-26 — identification of a pathogenic variant in *RNU4-2*

A female patient in the 16–20 age range presented with severe developmental delay, intellectual disability, microcephaly (head circumference −5.18 SDS), autism spectrum features, self-mutilating behavior, and hydronephrosis. Supported sitting was achieved in late childhood and walking in early adolescence with physiotherapy. No meaningful speech was present. Previous karyotyping, array CGH, and WES were non-diagnostic.

RNA-seq–based variant calling identified a heterozygous n.64_65insT variant in *RNU4-2* (NR_003137.3). (Figure 6b) The finding was confirmed by Sanger sequencing and targeted validation, and parental testing established the variant as de novo. (Figure 6b,c). This variant is the most frequently reported pathogenic variant associated with ReNU syndrome[65, 66] and disrupts the interaction between U4 and U6 snRNAs, impairing spliceosomal catalytic activation and causing widespread splicing dysregulation. This case illustrates the critical role of RNA-seq in detecting pathogenic variants in non-protein-coding RNA genes, which are typically not targeted or reliably analyzed by standard WES pipelines.

#### Overall diagnostic yield across groups

Across the entire cohort, RNA-seq contributed to a molecular diagnosis in 29 of 88 evaluable patients (32.9%). The diagnostic yield differed substantially across the three patient subgroups: 82.8% (24/29) in Group 1 (hypothesis-driven splicing analysis of candidate VUS), 6.9% (2/29) in Group 2 (patients with a specific pre-diagnosis but no identified pathogenic variant), and 10% (3/30) in Group 3 (patients without a pre-diagnosis or candidate variant). These results demonstrate that RNA-seq has the highest diagnostic utility when applied to evaluate specific candidate variants with predicted splicing impact, while its yield as an unbiased screening tool in the absence of a candidate variant is substantially lower but not negligible.

## Discussion

In this study, we systematically evaluated the diagnostic utility of RNA-seq across three molecularly and phenotypically stratified subgroups of patients with non-diagnostic exome sequencing. The overall diagnostic yield was 32.9%, but this aggregate figure masked a pronounced gradient across groups, with hypothesis-driven analysis of candidate splice variants substantially outperforming unbiased transcriptomic screening. These findings demonstrate that the diagnostic contribution of RNA-seq is highly dependent on cohort composition and patient selection, with the greatest utility observed in the functional validation of candidate splice variants identified by prior DNA-based testing.

Our overall diagnostic yield was higher than the range previously reported in blood-based heterogeneous RNA-seq cohorts (2.6–22.5%).[12, 15, 17, 18, 21–23]. This relatively high yield is largely attributable to the enrichment of Group 1 patients with candidate splice variants. The diagnostic rate of 82.8% in this subgroup is consistent with previous studies demonstrating that hypothesis-driven RNA-seq analysis of candidate variants achieves substantially higher yields than unbiased transcriptomic screening.[13, 21] Conversely, our yields of 6.9% in Group 2 and 10% in Group 3 are comparable to those reported in studies employing RNA-seq without prior candidate variants, where diagnostic rates of 1.5–21% have been described [6, 7, 10, 11, 16, 17, 21].

Several strategies contributed to the diagnostic success in Groups 2 and 3 despite the absence of candidate variants. First, a phenotype-driven approach was adopted, using clinically informed gene lists to prioritize findings from the large number of statistical outliers generated by RNA-seq analyses. This strategy proved critical in the case of G2-2, where a well-established pathogenic *ACTB* variant present at a low allele fraction (∼11%) was identified through RNA-seq variant calling after being missed by WES due to its somatic mosaic nature. This case illustrates the value of integrating clinical phenotyping with molecular data, rather than relying solely on bioinformatic thresholds. Second, the inclusion of affected family members, particularly siblings, strengthened the analytic power of RNA-seq. In Group 3, the identification of shared RNA aberrations in sibling pairs (G3-14/15) was instrumental in establishing a diagnosis and reduced the likelihood of incidental findings, underscoring the value of family-based approaches in RNA-seq studies.

A key factor contributing to the lower diagnostic yield in Groups 2 and 3 is the inherent limitation of blood as a tissue source for RNA-seq. In our cohort, the median number of protein-coding genes detectably expressed in peripheral blood was 11,652 at TPM > 1 (range, 7,014–12,100) and 12,419 at TPM > 0.5 (range, 9,130–13,009), indicating that a substantial fraction of the protein-coding transcriptome falls below thresholds required for confident splicing or expression outlier analysis. Moreover, adequate gene-level expression does not guarantee informative splicing analysis. In two patients with suspected neurofibromatosis type 1 (G2-3 and G2-15), although *NF1* appeared sufficiently expressed in blood based on GTEx Portal v10 release (median TPM = 1.87), the majority of reads mapped to a shorter, non-canonical transcript (ENST00000689464.1) rather than to the clinically relevant canonical isoform, resulting in insufficient read coverage at critical exons. A similar limitation was observed for *NPHP4* in patient G1-3: although gene-level expression appeared acceptable according to GTEx (median TPM = 2.08), the canonical transcript (ENST00000378156.9) had a substantially lower expression in the same reference dataset (median TPM = 0.24), and the candidate variant region could not be reliably assessed. These observations suggest that patient selection for blood-based RNA-seq should rely not only on gene-level expression in reference databases such as GTEx, but also on transcript-, exon-, and splice junction–level coverage of the clinically relevant isoform.

A counterpoint to these expression-limited cases is provided by the somatic mosaic *ACTB* variant identified in G2-2. As a highly expressed structural gene (TPM = 3,559), *ACTB* yielded sufficient read depth (96,899 reads at the variant position) to confidently detect the variant despite its low allele fraction (∼11%), a level of sensitivity that would be unattainable for the lowly expressed transcripts described above. Together, these observations indicate that expression level governs variant detectability at both extremes: insufficient coverage precludes assessment in poorly expressed genes, whereas abundant expression enables reliable detection of even low-level mosaic events.

The identification of a pathogenic *RNU4-2* variant in case G3-26 illustrates that snRNA loci can be interrogated by blood-based RNA-seq, but their analytical accessibility is gene-specific and not guaranteed across the cohort. To quantify this, we examined gene-level expression of the four snRNA genes manually inspected in patients with neurodevelopmental phenotypes (*RNU2-2*, *RNU4-2*, *RNU5A-1*, and *RNU5B-1*) across the 87 QC-passing samples. Median expression levels varied by approximately an order of magnitude across loci (median TPM: *RNU5A-1* 39.4; *RNU2-2* 9.9; *RNU4-2* 6.1; *RNU5B-1* 1.9), with corresponding differences in the proportion of samples reaching an analytically informative threshold of TPM ≥ 1: *RNU5A-1* in 98.9% (86/87), *RNU2-2* in 93.1% (81/87), *RNU4-2* in 89.7% (78/87), and *RNU5B-1* in only 73.6% (64/87) of samples. For *RNU4-2* specifically — the snRNA gene that yielded a diagnosis in this cohort — 7/87 samples (8.0%) showed no detectable expression (TPM = 0), and a further 2 samples fell below TPM 1, indicating that approximately 10% of patients would have been analytically uninformative for *RNU4-2* manual inspection had they harbored a relevant variant.

This sample-level variability was partly driven by sequencing depth. Across the 87 QC-passing samples, snRNA raw read counts scaled sub-linearly with total post-trimming read count on the log-log scale, with regression slopes of 0.84 for *RNU2-2*, 0.74 for *RNU4-2*, 0.61 for *RNU5A-1*, and 0.28 for *RNU5B-1* (Supplementary Figure S25). On the untransformed scale, Pearson correlations between library size and snRNA recovery were weak (|r| ≤ 0.12, p ≥ 0.27), reflecting the influence of a small number of high-recovery outliers, while Spearman correlations were modest but significant for three of the four loci (r = 0.21–0.28, p = 0.009–0.055). Together, these observations indicate that library size contributes systematically but accounts for only a small fraction of the inter-sample variance in snRNA recovery. A substantial residual component was sample-specific rather than locus-specific: the four snRNA genes were strongly correlated with one another at the sample level (raw-read Pearson r: *RNU2-2*–*RNU4-2* = 0.93; *RNU5A-1*–*RNU5B-1* = 0.88; all between-locus comparisons r ≥ 0.75), pointing to a shared, sample-specific source of snRNA capture in poly-A–selected libraries whose mechanistic basis is uncertain and may involve stochastic oligo(dT) capture of partially adenylated pre-snRNA intermediates together with unmeasured pre-analytical or library preparation variables. Regardless of mechanism, these observations indicate that, although ReNU syndrome and other snRNA-associated disorders can be diagnosed from peripheral blood RNA-seq, snRNA recovery from poly-A–selected libraries is variable in a manner not fully predictable from library size alone, and dedicated total-RNA or small-RNA protocols would be required for systematic, predictable coverage of this gene class.

The other key factor contributing to the lower diagnostic yield is the limited performance of aberrant splicing detection tools. FRASER and dasper produced largely non-overlapping per-sample call sets in our cohort: FRASER detected a median of 211 outlier genes per sample (range: 0–465) compared with 164 by dasper (range: 0–455), with only a median of 24 genes (range: 0–61) detected concordantly by both tools. Of the FRASER calls, a median of 10.8% per sample (range: 2.4–33.3%) were also flagged by dasper, while a median of 14.3% of dasper calls per sample (range: 0.2–31.1%) were captured by FRASER. This asymmetric and partial overlap reflects the methodological distinction between the two approaches: FRASER models aberrant splice-junction usage through a Beta-binomial framework operating on intron-centric splice metrics, while dasper applies an outlier-detection framework over splice-junction read patterns ranked against a reference cohort.

Although such partial overlap might be expected to indicate complementary detection of distinct aberrant events, this was not reflected in our diagnostic outcomes: FRASER independently prioritized no cases beyond those already detected by dasper. Among the Group 1 patients with confirmed aberrant splicing, dasper identified 15 of 24 events, while FRASER identified only 5 — all of which were also captured by dasper. In Groups 2 and 3, the *IGBP1* deep intronic event in G3-14/15 was detected by dasper but missed by FRASER, while the *APPL1* event in G2-7 was missed by both transcriptome-wide splicing outlier tools and was instead resolved through SpliceAI-guided variant-level review. These findings are consistent with previous reports: De Cock et al. observed detection of 4 of 6 positive cases by FRASER[67], and Jaramillo Oquendo et al. — using the more recent FRASER2[68] algorithm, which has been reported to offer improved sensitivity over the original FRASER — detected only 4 of 14 IGV-confirmed aberrant splicing events [17]. Tools that struggle to reliably detect confirmed aberrant splicing events cannot be expected to perform well in the unbiased discovery setting required for Groups 2 and 3, which partially explains the lower diagnostic yield in these subgroups.

In contrast, the *in silico* splicing prediction tool SpliceAI demonstrated 54.1% full and 29.1% partial concordance with observed splicing events in Group 1. Furthermore, SpliceAI predictions guided the prioritization of the *APPL1* variant in G2-7 and correctly predicted the effect of the *IGBP1* variant in G3-14/15. These observations support the use of SpliceAI as a reliable variant triage tool in the RNA-seq workflow. However, it is important to recognize that variants creating new alternative splice sites without abolishing the canonical site often produce’leaky’ splicing, in which a proportion of transcripts are processed normally. In such cases, even variants with high SpliceAI scores may yield limited aberrant signal in RNA-seq, particularly when the target gene shows tissue-restricted expression. This is illustrated by case G1-4, in which a CTLA4 synonymous variant (c.243G>A) with a donor gain prediction (donor gain [DG]: 0.26) — but no predicted loss of the canonical donor — showed only minimal aberrant donor usage in blood-derived RNA (1/55 reads) (Additional file 3: Supplementary figure S24). Given that CTLA4 is most highly expressed in activated T cells and regulatory T cells rather than the predominant cell types in whole blood, the modest signal may be consistent with both the leaky splicing scenario and tissue underrepresentation. The variant remained classified as VUS (PM2, PP3), and stimulated T-cell or sorted Treg RNA analysis would be required for definitive functional assessment. Such cases underscore that a negative or modest blood RNA-seq result does not exclude clinically relevant aberrant splicing in target tissues and should inform both patient selection and variant interpretation.

Expression outlier analysis had limited diagnostic utility in this cohort. The detection of numerous outlier genes per patient reduced specificity, and for disorders primarily manifesting in tissues other than blood, expression-level assessment is inherently constrained. Notably, many patients harboring heterozygous variants predicted to trigger NMD did not show significant reductions in gene expression, a finding consistent with the persistence of wild-type allele expression in the heterozygous state and additionally influenced by tissue-specific NMD efficiency and technical noise. We therefore consider aberrant expression as supportive rather than independently diagnostic evidence in blood-based RNA-seq. Normalization against housekeeping genes may further improve its diagnostic performance.

A further methodological limitation of short-read RNA-seq is its inability to resolve the full structure of aberrant transcripts when multiple events occur within the same gene or when transcripts terminate at non-canonical positions. Two cases in our cohort illustrate this limitation. In proband G1-20, short-read RNA-seq detected exon 4 skipping concurrent with retention of introns 3 and 4 but could not determine whether these events co-occurred within a single aberrant transcript or arose from independent transcripts, precluding unambiguous prediction of the resulting protein consequence (Additional file 3: Supplementary figure S14). In proband G1-18, an aberrant junction connecting the canonical 5′ donor of exon 7 to a cryptic acceptor within the terminal exon of an annotated short isoform was supported by 26 reads, with no further junctions extending to downstream canonical exons (Additional file 3: Supplementary figure S12). This pattern most likely reflects alternative last exon usage involving the short isoform, although short-read data alone cannot formally exclude additional downstream splicing events.

In both cases, definitive transcript-level interpretation would require long-read RNA-seq, which captures full-length transcripts and enables direct phasing of splicing events. The integration of long-read RNA-seq into clinical diagnostic workflows, although currently constrained by cost and lower per-base accuracy compared with short-read platforms, is likely to become increasingly relevant for resolving such cases.

This study has several strengths, including the systematic stratification of patients into three molecularly and phenotypically distinct groups, enabling direct comparison of RNA-seq utility across different clinical scenarios. The integration of phenotype-driven prioritization, family-based analysis, and comparative evaluation of multiple bioinformatic tools enhanced the analytic rigor. However, important limitations should be acknowledged. No NMD inhibitors were used, potentially reducing the detection sensitivity for variants producing transcripts subject to rapid degradation. This limitation may decrease the apparent fraction of aberrant transcripts and complicate interpretation, as illustrated by cases G1-26, G1-30 and G2-7 (Additional file 3: Supplementary Figures S19, S21, S22). The use of GTEx as a reference cohort, rather than an age-matched internal cohort processed with identical protocols, may have introduced confounding variation. Finally, the majority of patients had undergone WES rather than WGS as their initial DNA test, which may have limited the ability to identify explanatory intronic or regulatory variants underlying some of the RNA-level aberrations observed.

## Conclusions

RNA-seq achieved a high diagnostic yield (82.8%) when applied to evaluate candidate VUS with predicted splicing impact, confirming its value as a powerful functional validation tool in clinical genetics. In patients without candidate variants, diagnostic yields were lower (6.9–10%) but not negligible, with RNA-seq enabling the identification of somatic mosaicism, deep intronic pseudoexon-activating variants, and pathogenic changes in non-protein-coding RNA genes that were undetectable by standard exome sequencing. The systematic comparison across three molecularly stratified patient groups demonstrated that RNA-seq performance is highly dependent on clinical context and patient selection. These findings support the integration of RNA-seq into clinical genomic workflows as a second-tier test following non-diagnostic DNA-based sequencing.

## List of abbreviations

ACGS: Association for Clinical Genomic Science
ACMG/AMP: American College of Medical Genetics and Genomics / Association for Molecular Pathology
AG: acceptor gain (SpliceAI)
AL: acceptor loss (SpliceAI)
ASE: allele-specific expression
BMI: body mass index
cDNA: complementary DNA
CGH: comparative genomic hybridization
ClinGen: Clinical Genome Resource
DG: donor gain (SpliceAI)
DL: donor loss (SpliceAI)
DTR: deep tendon reflex
EDTA: ethylenediaminetetraacetic acid
EEG: electroencephalography
EMG: electromyography
FRASER: Find RAre Splicing Events in RNA-seq
GATK: Genome Analysis Toolkit
GTEx: Genotype-Tissue Expression
HJS: Houge–Janssens syndrome
HPO: Human Phenotype Ontology
IFM: isolation forest model
IGV: Integrative Genomics Viewer
MAE: monoallelic expression
MAF: minor allele frequency
MRI: magnetic resonance imaging
NDD/ID: neurodevelopmental disorder / intellectual disability
NGS: next-generation sequencing
NMD: nonsense-mediated decay
OMIM: Online Mendelian Inheritance in Man
PAE: predicted aligned error
PCA: principal component analysis
PCR: polymerase chain reaction
pLDDT: predicted local distance difference test
PVS1_RNA: PVS1 evidence based on RNA-seq–demonstrated loss of function
QC: quality control
RIN: RNA integrity number
RNA-seq: RNA sequencing
SNP: single-nucleotide polymorphism
snRNA: small nuclear RNA
SPRI: solid-phase reversible immobilization
STAR: Spliced Transcripts Alignment to a Reference
SVI: Sequence Variant Interpretation
TMM: trimmed mean of M-values
TPM: transcripts per million
VEP: Variant Effect Predictor (Ensembl); visual evoked potential (clinical context)
VUS: variant of uncertain significance
WES: whole-exome sequencing
WGS: whole-genome sequencing

## Declarations

### Ethics approval and consent to participate

This study was approved by the Clinical Research Ethics Committee of Gazi University Faculty of Medicine (decision dated December 6, 2024; approval number 42). Written informed consent was obtained from all participants or their legal guardians prior to inclusion in the study, in accordance with the Declaration of Helsinki.

### Consent for publication

Written informed consent for publication of clinical details and clinical images was obtained from the patient or, where the patient was a minor, from the parent or legal guardian. Copies of the consent forms are available for review by the Editor-in-Chief on request.

### Availability of data and materials

The RNA-seq datasets generated and analyzed during the current study contain potentially identifying genetic and clinical information from patients with rare Mendelian disorders. In accordance with the conditions of ethical approval (Clinical Research Ethics Committee of Gazi University Faculty of Medicine; decision dated December 6, 2024; approval number 42) and the informed consent obtained from participants and their legal guardians, raw sequencing data cannot be deposited in a public repository. Processed and de-identified data supporting the findings of this study, including variant-level results and summary expression and splicing metrics, are available from the corresponding author on reasonable request, subject to a data access agreement and approval by the Gazi University Faculty of Medicine Clinical Research Ethics Committee. Reference RNA-seq data from the Genotype-Tissue Expression (GTEx) Consortium v8 release used as the control cohort are publicly available at https://gtexportal.org/.

### Competing interests

S.D., F.E.S., and T.C. are co-founders, shareholders, and employees of PhiTech, a company specializing in RNA sequencing analysis pipelines. The remaining authors declare no competing interests.

### Funding

This study was supported by PhiTech, which funded RNA sequencing library preparation, sequencing, and bioinformatic analyses. Three authors (S.D., F.E.S., T.C.) are affiliated with PhiTech (see Competing interests).

### Authors’ contributions

T.D., G.K., and M.A.E. conceived and designed the study. T.D., H.E.K., G.K., and M.A.E. recruited patients, collected clinical data. S.D., F.E.S., and T.C. designed and implemented the bioinformatic analysis pipeline and performed all bioinformatic analyses. T.D., H.E.K and M.A.E. analyzed the data and interpreted variants. T.D. wrote the first draft of the manuscript. All authors critically reviewed the manuscript and approved the final version.

## Data Availability

The RNA-seq datasets generated and analyzed during the current study contain potentially identifying genetic and clinical information from patients with rare Mendelian disorders. In accordance with the conditions of ethical approval (Clinical Research Ethics Committee of Gazi University Faculty of Medicine; decision dated December 6, 2024; approval number 42) and the informed consent obtained from participants and their legal guardians, raw sequencing data cannot be deposited in a public repository. Processed and de-identified data supporting the findings of this study, including variant-level results and summary expression and splicing metrics, are available from the corresponding author on reasonable request, subject to a data access agreement and approval by the Gazi University Faculty of Medicine Clinical Research Ethics Committee.

## Acknowledgements

We are deeply grateful to the patients and their families for their participation in this study and for their trust in our research. We thank Elif Gündüz, Cihangir Taşkan and Oğuz Eröz for their excellent technical assistance throughout the project. We are also grateful to Dr. Ozan Vural, Dr. Hacer Demet Özcan, Dr. Ali Babazade, Dr. Esra Güneş, Dr. Ekin Alpaslan, Dr. Tilbe Hakçıl, and Dr. Yusuf Bahap for their valuable clinical contributions and patient referrals. We acknowledge the Genotype-Tissue Expression (GTEx) Project for providing the reference transcriptomic data used in this study.

## Additional files

**Additional file 1: Supplementary Methods.** Detailed protocols for fibroblast culture and DNA extraction (patient G2-2), structural prediction of wildtype and aberrantly spliced *RB1* protein isoforms using ColabFold (patient G1-29), and primer sequences used for variant confirmation by Sanger sequencing and targeted next-generation sequencing (Table SM1).

**Additional file 2: Tables S1–S3.** Clinical summaries and molecular findings for all enrolled patients, stratified by group (Table S1, Group 1; Table S2, Group 2; Table S3, Group 3).

**Additional file 3: Supplementary Figures S1–S25.** Sashimi plots and IGV alignments of RNA-seq findings, including positive cases not highlighted in the main text. Selected examples illustrating methodological limitations of blood-based short-read RNA-seq. Cohort-level analysis of the relationship between sequencing depth and snRNA gene expression.

## Notes

### Summary of Updates

1)Added a new paragraph in the Discussion contrasting the ACTB somatic mosaicism case (G2-2) with the expression-limited cases (NF1, NPHP4, CTLA4), highlighting that high transcript abundance (whole-blood median TPM = 3,559) enabled confident detection of the variant despite its low (~111% allele fraction. 2) Specified the GTEx data release used for gene-specific expression values cited in the text (GTEx Portal v10) in the Discussion. 3) Revised the Discussion statement on heterozygous NMD-predicted variants: removed the reference to intron retention artifactually inflating expression estimates and replaced it with persistence of wild-type allele expression as the primary explanation, alongside tissue-specific NMD efficiency and technical noise. 4) Corrected panel labels within Figures 4 and 5. 5) Repositioned figures and Table 1 from the end of the manuscript to their corresponding locations within the main text.

