## Additional file 1-Supplementary_Methods for "Stratified evaluation of blood RNA sequencing in a rare disease cohort"

### Primer sequences for variant confirmation

Primers flanking the target regions were designed using NCBI Primer-BLAST. Sequences used for variant confirmation by Sanger sequencing and targeted next-generation sequencing are listed in Table SM1.

**Table SM1.** Primer sequences used for variant confirmation by Sanger sequencing and targeted next-generation sequencing.

| Patient ID | Gene | Variant | Forward primer (5′→3′) | Reverse primer (5′→3′) | Amplicon size (bp) | Method |
|---|---|---|---|---|---|---|
| G1-29 (proband, parents) | *RB1* | NM_000321.3: c.1332+4A>G | GTCTGCTTATGTTCAGTAGTTGTGG | CCCATAAATAGCAGCATACACAGG | 431 | Sanger |
| G2-2 | *ACTB* | NM_001101.5: c.359C>T (p.Thr120Ile) | GAGAAAATCTGGCACCACACC | CACCTAGTCAGAGAGACAAACACC | 319 | Sanger + targeted NGS |
| G3-14, G3-15 | *IGBP1* | NM_001551.3: c.872-3371T>A | ATACCACTTTGGTCAGGTCATCG | GTACTGAGTTCCTACATAACCACACC | 438 | Sanger |
| G3-26 (proband, parents) | *RNU4-2* | NR_003137.3: n.64_65insT | ATGCAGATGGGCCTTAAATACG | AGTGACAAAGGAAGTGGAGTAGC | 559 | Sanger + targeted NGS |

### Fibroblast culture and DNA extraction (patient G2-2)

A 4-mm punch skin biopsy was obtained from the medial aspect of the upper arm under local anesthesia after written informed consent. The biopsy was transferred to the laboratory in sterile transport medium and processed within 24 hours. The tissue was minced into small fragments and plated in T-25 culture flasks containing AmnioMAX™-II Complete Medium (Gibco, Thermo Fisher Scientific). Cultures were maintained at 37 °C in a humidified atmosphere with 5% $CO_2$. Medium was refreshed every 3–4 days until sufficient fibroblast outgrowth was obtained. Genomic DNA was extracted directly from primary cultures without further passaging, using the QIAamp DNA Mini Kit (Qiagen, Valencia, CA) according to the manufacturer’s instructions.

**Structural prediction of wildtype and aberrantly spliced protein isoforms (patient G1-29)**

Three-dimensional structures of the wildtype and aberrantly spliced *RB1* isoforms were predicted using ColabFold v1.5.5, which combines MMseqs2-based homology search with AlphaFold2. Default parameters were used (3 recycles, 5 models per prediction, AlphaFold2-ptm). The unrelaxed top-ranked model (highest mean pLDDT) was selected for visualization, and per-residue confidence (pLDDT) and predicted aligned error (PAE) plots were inspected for all models. The mutant sequence was generated by deleting the residues encoded by the skipped exon (p.(Asn406_Gln444del)) from the canonical UniProt sequence (P06400). Structures were rendered in PyMOL v3 (Open Source) with functional domains annotated according to UniProt feature annotations.
