## Additional file 3-Supplementary figures for "Stratified evaluation of blood RNA sequencing in a rare disease cohort"

### Supplementary Figures S1–S25: RNA-seq findings

This document contains visualizations of all RNA-seq findings from positive and selected negative cases not highlighted in the main text, together with cohort-level analyses supporting the interpretation of snRNA gene coverage. Captions provide patient and variant details, the observed splicing aberration, and predicted RNA and protein consequences.

#### Supplementary Figures S1–S22 — Positive cases

Sashimi plots of the 22 positive RNA-seq cases not highlighted in the main text. For each case, the proband's read alignments at the affected locus are shown in red and an unrelated inhouse control sample is shown in blue. Each panel demonstrates the splicing aberration attributable to the candidate variant and contrasts it with canonical splicing observed in the control sample.

#### Supplementary Figures S23–S24 — Examples of methodological limitations

Two cases from Group 1 in which the candidate variant could not be RNA-confirmed, illustrating distinct methodological limitations of blood-based short-read RNA-seq, shown as Integrative Genomics Viewer (IGV) alignment views of the proband. Supplementary Figure S23 (*NPHP4*, proband G1-3) shows insufficient read coverage at the candidate locus, precluding reliable assessment of aberrant splicing. Supplementary Figure S24 (*CTLA4*, proband G1-4) shows a SpliceAI-predicted aberrant splicing event detected at minimal read support, indistinguishable from technical noise. In both cases, the candidate variant was retained as of uncertain significance.

#### Supplementary Figure S25 — Library size and snRNA expression

Cohort-level analysis of the relationship between sequencing depth and snRNA gene expression across the 87 QC-passing samples.

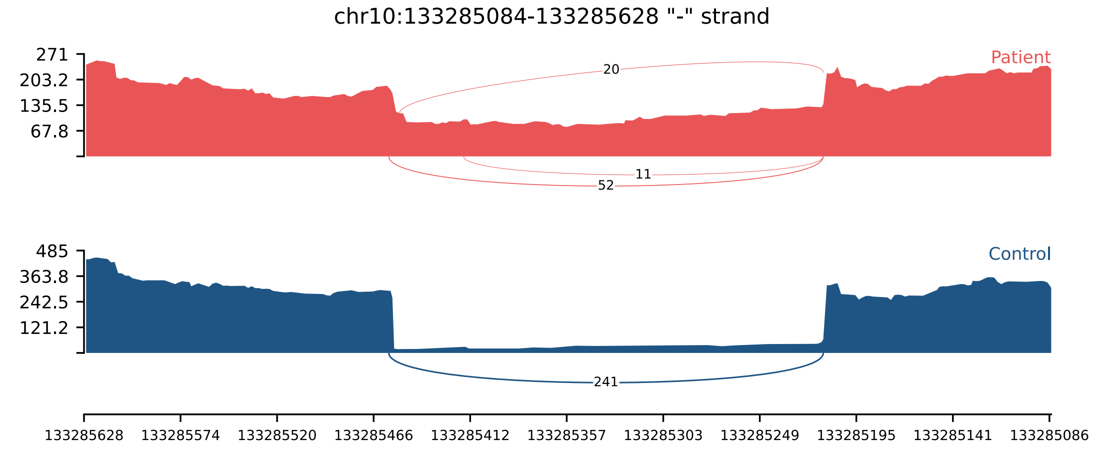


**Supplementary Figure S1:** Sashimi plot of homozygous *TUBGCP2* (NM_006659.4):c.1895+5G>A variant. RNA-seq in proband G1-2 (red) demonstrates intron retention and 6-bp or 44-bp exon extension due to alternative donor site usage in contrast to minimal intron retention observed in unrelated inhouse control sample (blue). The aberrant splicing produces three transcripts (r.[1895_1896ins1895+1_1895+6, 1895_1896ins1895+1_1895+44, 1895_1896ins1895+1_1896-1]), predicted to result in p.[(Arg632_Lys633insCysGly), (Lys633Cysfs*15), (Lys633Cysfs*15)], respectively.

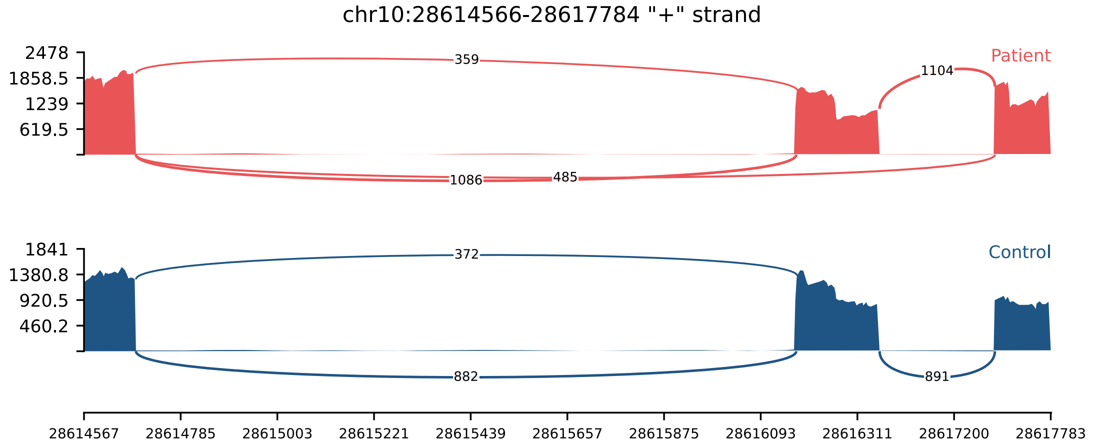


**Supplementary Figure S2:** Sashimi plot of heterozygous *WAC* (NM_016628.5):c.1746+3A>G variant. RNA-seq in proband G1-5 (red) demonstrates exon skipping in contrast to normal splicing observed in unrelated inhouse control sample (blue). The aberrant transcript (r.1557_1746del) is predicted to result in p.(Ser519Argfs*24).

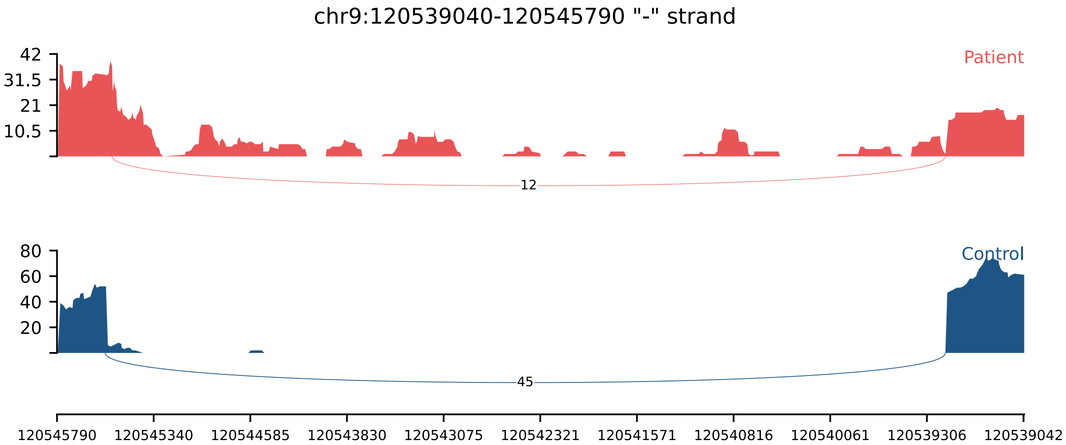


**Supplementary Figure S3:** Sashimi plot of homozygous *CDK5RAP2* (NM_018249.6):c.383+4dup variant. RNA-seq in proband G1-6 (red) demonstrates 49-bp exon extension due to alternative donor site usage and intron retention in contrast to normal splicing observed in unrelated inhouse control sample (blue). The aberrant splicing produces two transcripts (r.[383_384ins383+1_383+49, 383_384ins383+1_384-1]), both predicted to result in p.(Lys129Ter).

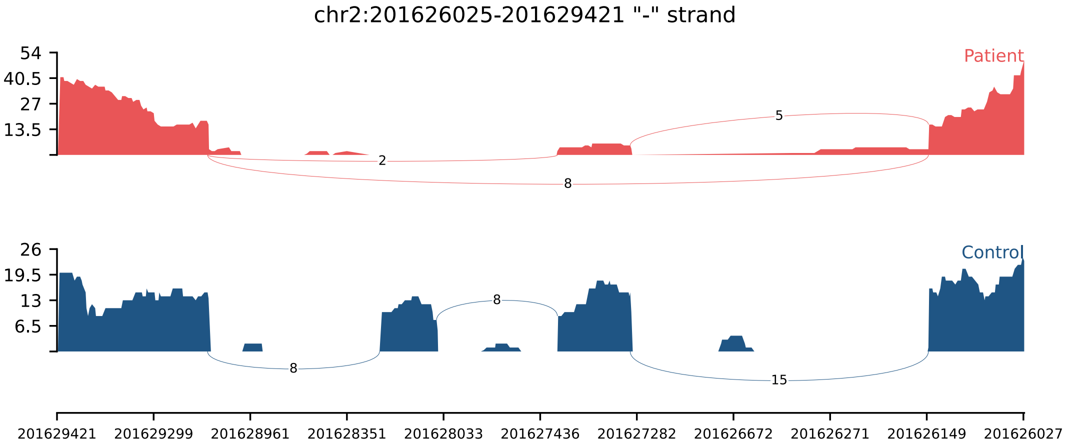


**Supplementary Figure S4:** Sashimi plot of homozygous *TMEM237* (NM_001044385.3):c.943+5G>A variant. RNA-seq in proband G1-7 (red) demonstrates skipping of two exons in contrast to normal splicing observed in unrelated inhouse control sample (blue). The aberrant transcript (r.870_1037del) is predicted to result in p.(Ile291_Trp346del).

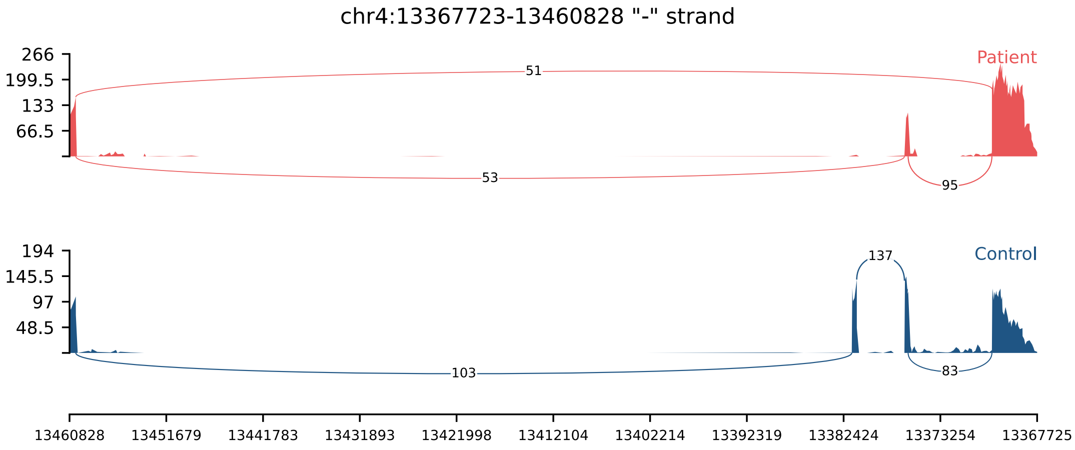


**Supplementary Figure S5:** Sashimi plot of homozygous *RAB28* (NM_001017979.3):c.495+6T>A variant. RNA-seq in proband G1-9 (red) demonstrates skipping of exon 5 in all reads, with concurrent skipping of exon 6 in approximately 50% of reads, in contrast to normal splicing observed in unrelated inhouse control sample (blue). The aberrant splicing produces two transcripts (r.[392_495del, 392_573del]), predicted to result in p.[(Ile131Serfs*11), (Ile131Lysfs*20)], respectively.

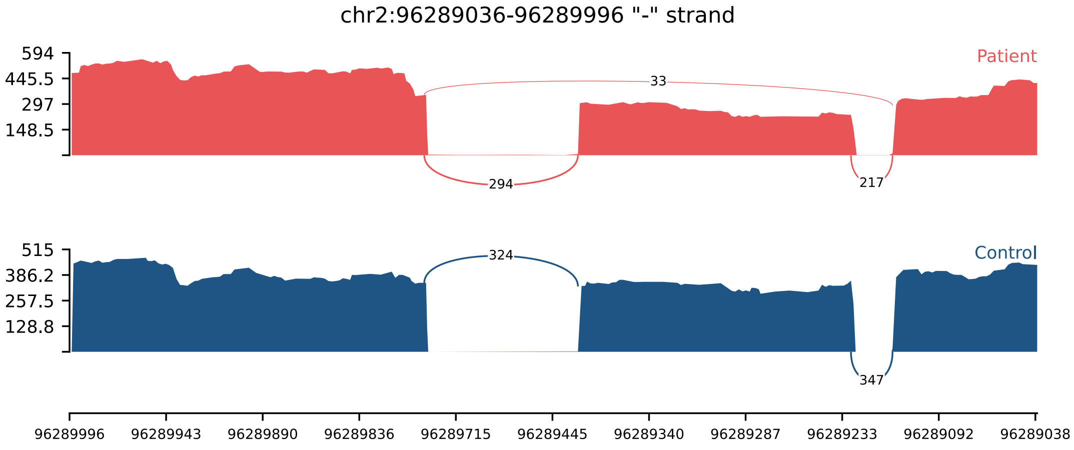


**Supplementary Figure S6:** Sashimi plot of heterozygous *SNRNP200* (NM_014014.5):c.3093G>A(p.Glu1031=) variant. RNA-seq in proband G1-10 (red) demonstrates exon skipping in contrast to normal splicing observed in unrelated inhouse control sample (blue). The aberrant transcript (r.2941_3093del) is predicted to result in p.(Val981_Glu1031del).

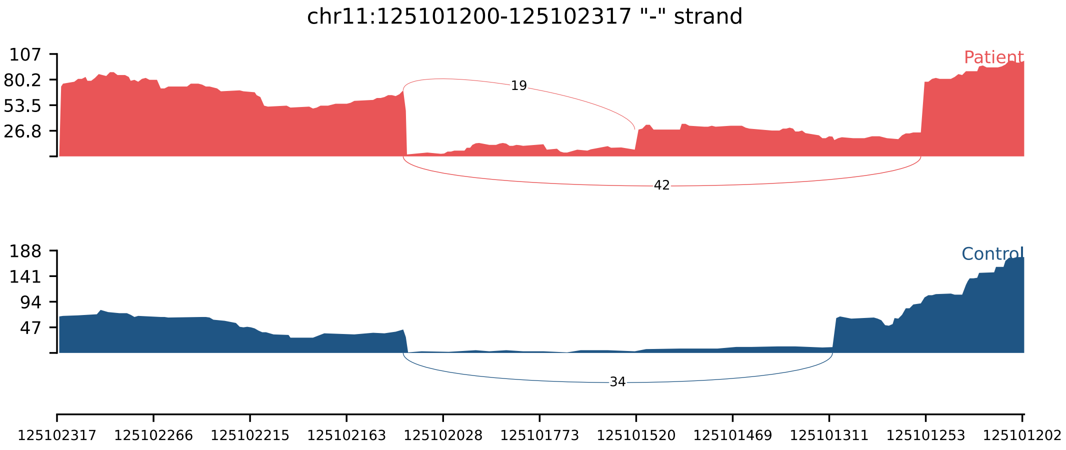


**Supplementary Figure S7:** Sashimi plot of homozygous *TMEM218* (NM_001258244.1):c.111G>T(p.Arg37Ser) variant. RNA-seq in proband G1-11 (red) demonstrates 47-bp exon shortening and 218-bp exon extension due to alternative acceptor site usage in contrast to normal splicing observed in unrelated inhouse control sample (blue). The aberrant splicing produces two transcripts (r.[111_157del, 110_111ins111-218_111-1]), predicted to result in p.[(Arg37Serfs*10), (Phe38Asnfs*39)], respectively.

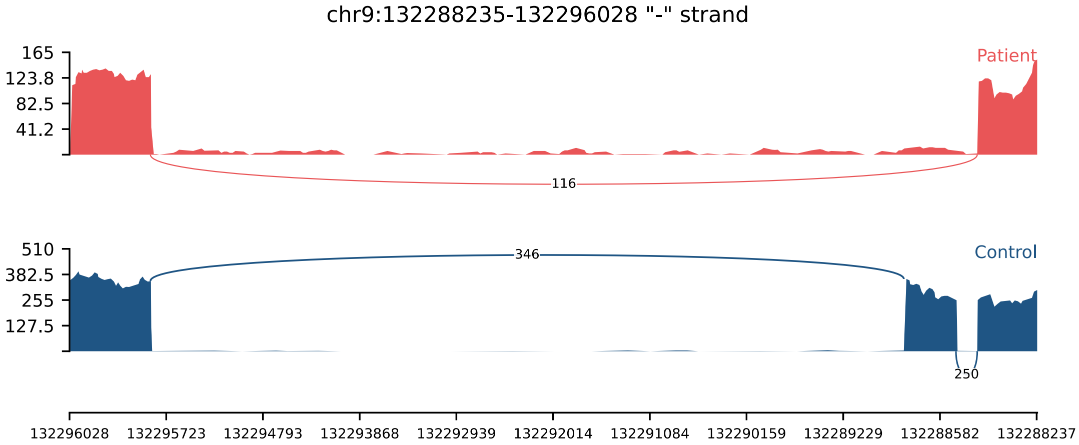


**Supplementary Figure S8:** Sashimi plot of homozygous *SETX* (NM_015046.7):c.6208+4A>G variant. RNA-seq in proband G1-13 (red) demonstrates exon skipping in contrast to normal splicing observed in unrelated inhouse control sample (blue). The aberrant transcript (r.6107_6208del) is predicted to result in p.(Gly2036_Lys2070delinsVal).

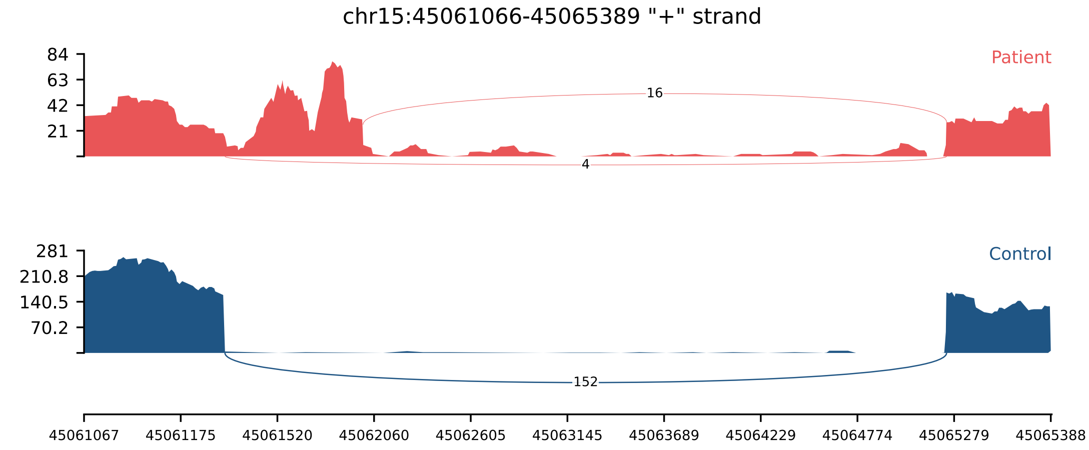


**Supplementary Figure S9:** Sashimi plot of homozygous *SORD* (NM_003104.6):c.425+6T>A variant. RNA-seq in proband G1-14 (red) demonstrates 771-bp exon extension due to alternative donor site usage in contrast to normal splicing observed in unrelated inhouse control sample (blue). The aberrant transcript (r.425_426ins425+1_425+771) is predicted to result in p.(Lys142_Leu143ins17*).

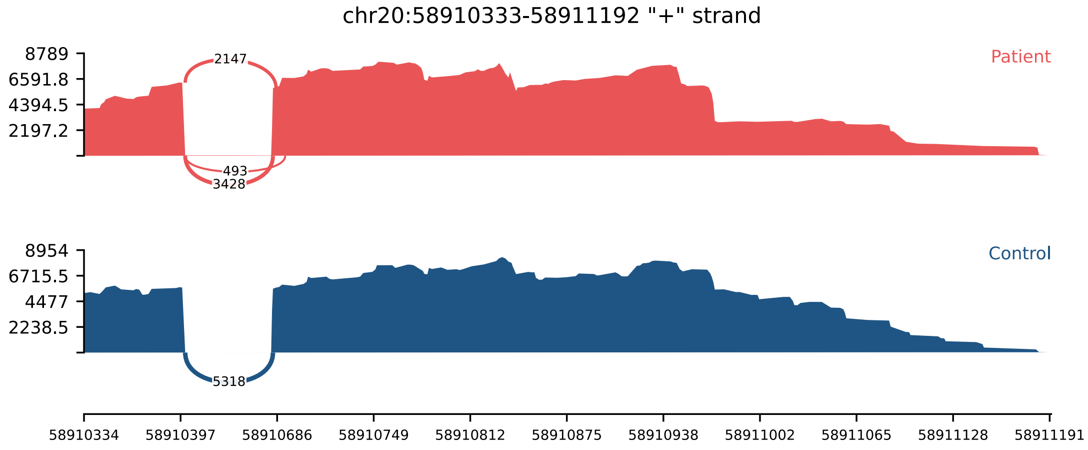


**Supplementary Figure S10:** Sashimi plot of heterozygous *GNAS* (NM_000516.7):c.1039-3C>G variant. RNA-seq in proband G1-15 (red) demonstrates 2-bp or 8-bp exon shortening due to alternative acceptor site usage in contrast to normal splicing observed in unrelated inhouse control sample (blue). The aberrant splicing produces two transcripts (r.[1039_1040del, 1039_1046del]), predicted to result in p.[(Arg347Aspfs*23), (Arg347Hisfs*21)], respectively.

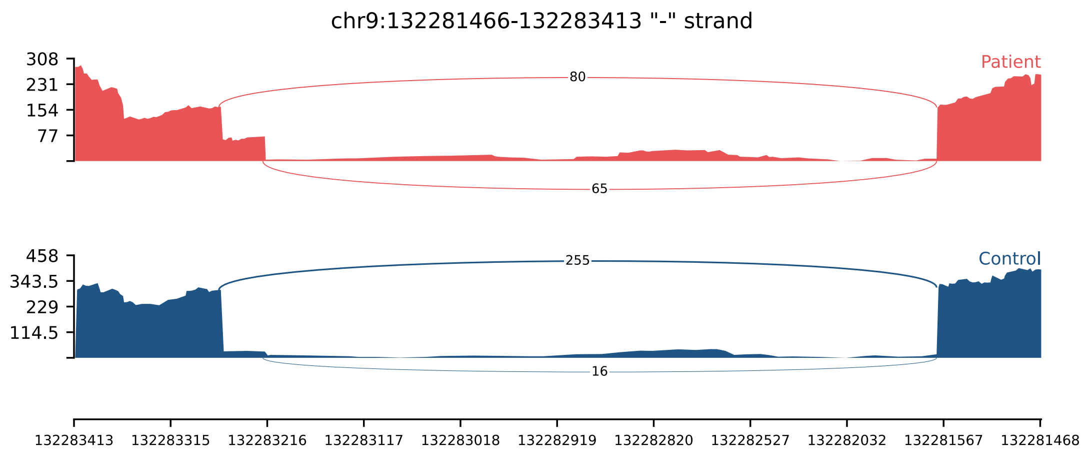


**Supplementary Figure S11:** Sashimi plot of heterozygous *SETX* (NM_015046.7):c.6546+5G>A variant. RNA-seq in proband G1-16 (red) demonstrates 45-bp exon extension due to alternative donor site usage in contrast to minimal exon extension observed in unrelated inhouse control sample (blue). The aberrant transcript (r.6546_6547ins6546+1_6546+45) is predicted to result in p.(Glu2182_Ala2183ins*10).

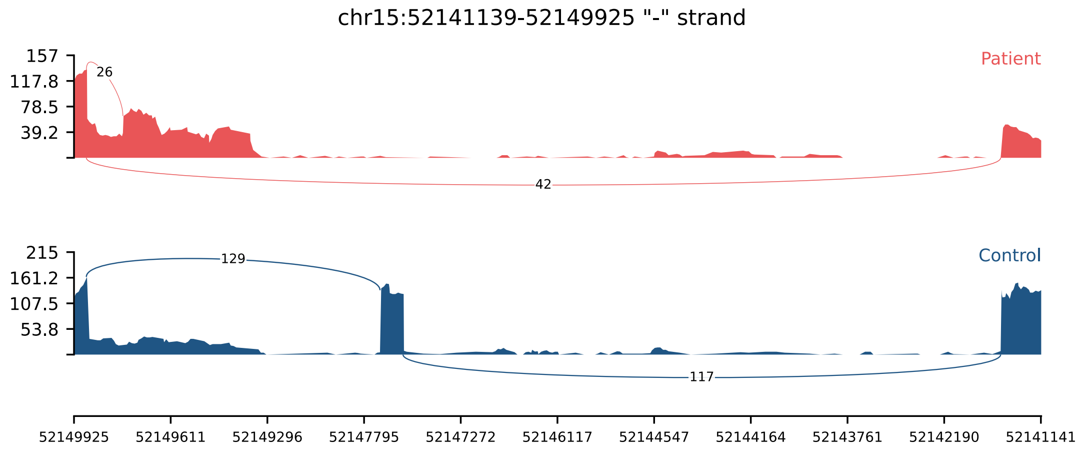


**Supplementary Figure S12:** Sashimi plot of homozygous *GNB5* (NM_016194.4):c.494+4A>G variant. RNA-seq in proband G1-18 (red) demonstrates two aberrant splicing events attributable to the variant, both absent or minimally observed in the unrelated in-house control sample (blue): (i) skipping of exon 7, yielding the transcript r.418_494del, predicted to result in p.(Glu140Trpfs*4); and (ii) usage of a cryptic acceptor site located within the terminal exon of the short isoform ENST00000560116.1, supported by 26 reads spanning the junction between the canonical 5′ donor of exon 7 and this cryptic acceptor (r.417_418ins417+118_417+606), predicted to introduce a premature termination codon (p.(Lys139_Glu140ins*15)). As no further junctions extending from this terminal exon back to downstream canonical exons were observed, this event may alternatively be interpreted as alternative last exon usage; however, definitive resolution would require long-read RNA-seq.

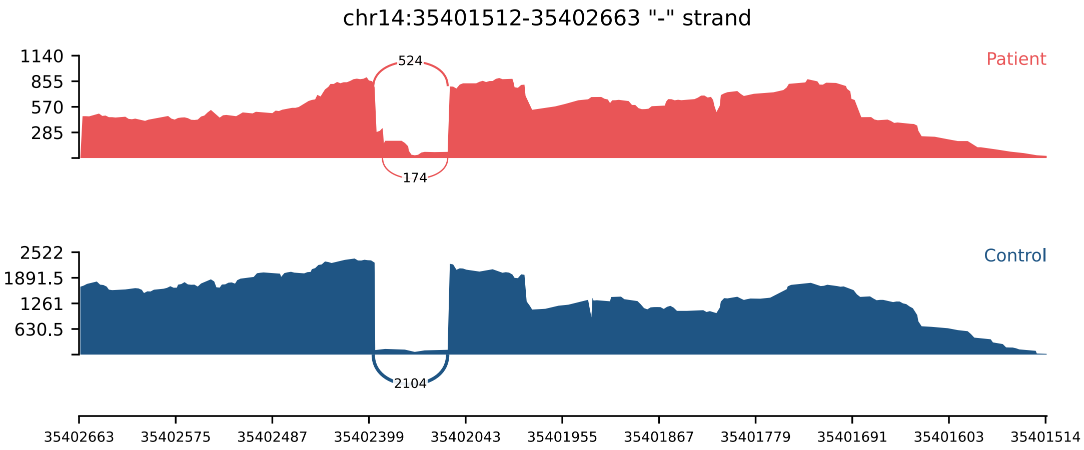


**Supplementary Figure S13:** Sashimi plot of heterozygous *NFKBIA* (NM_020529.3):c.906G>C(p.Glu302Asp) variant. RNA-seq in proband G1-19 (red) demonstrates 38-bp exon extension due to alternative donor site usage in contrast to normal splicing observed in unrelated inhouse control sample (blue). The aberrant transcript (r.906_907ins906+1_906+38) is predicted to result in p.(Leu303Valfs*4).

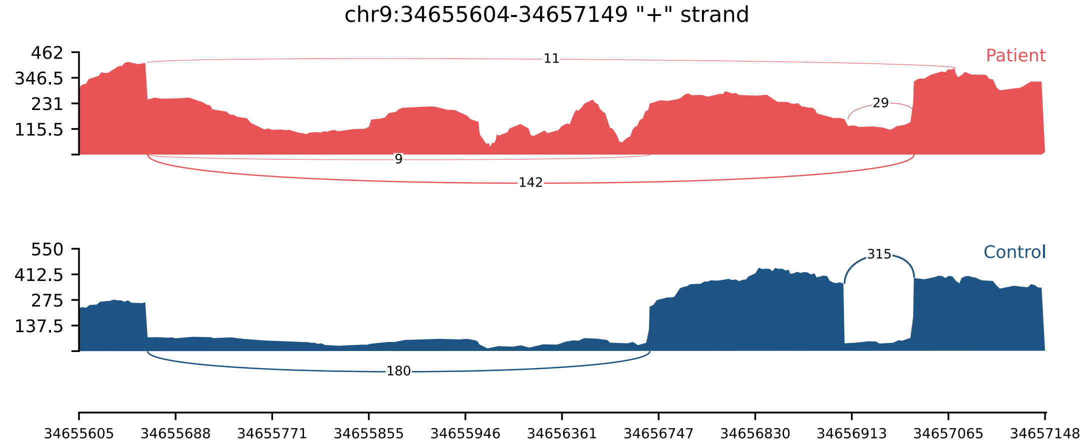


**Supplementary Figure S14:** Sashimi plot of homozygous *IL11RA* (NM_001142784.3):c.331+6T>G variant. RNA-seq in proband G1-20 (red) demonstrates exon 4 skipping together with retention of introns 3 and 4 in contrast to minimal retention of both introns observed in unrelated inhouse control sample (blue). The aberrant splicing yields an exon-skipped transcript (r.162_331del), predicted to result in p.(Asp55Profs*15), alongside intron retention events (r.161_162ins161+1_162-1 and r.331_332ins331+1_332-1) whose protein consequences cannot be unambiguously resolved by short-read RNA-seq, as the retention of introns 3 and 4 may occur in the same transcript or in independent transcripts.

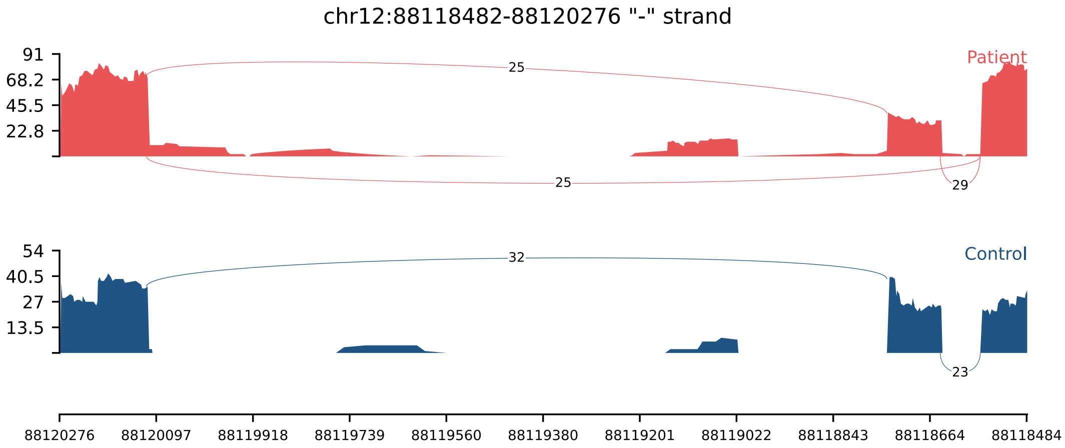


**Supplementary Figure S15:** Sashimi plot of heterozygous *CEP290* (NM_025114.3):c.1623+5G>C variant. RNA-seq in proband G1-21 (red) demonstrates exon skipping in contrast to normal splicing observed in unrelated inhouse control sample (blue). The aberrant transcript (r.1523_1623del) is predicted to result in p.(Gly508Aspfs*2).

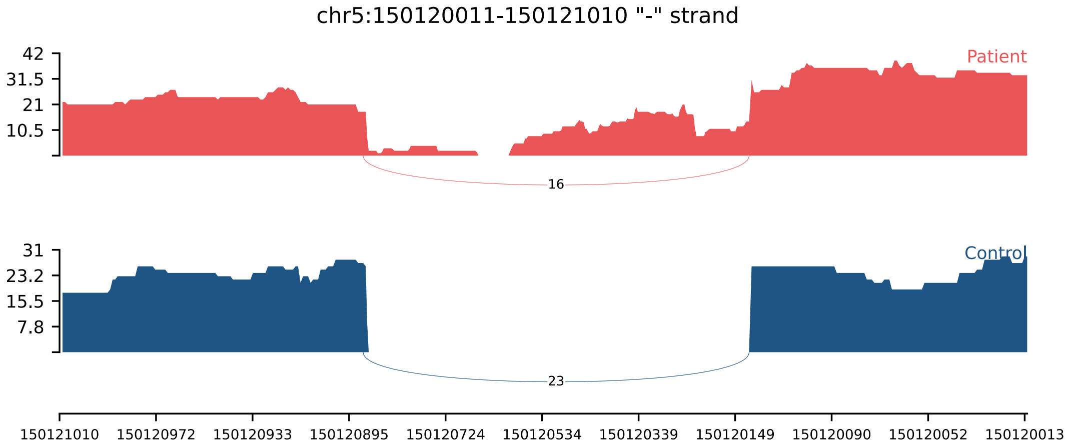


**Supplementary Figure S16:** Sashimi plot of heterozygous *PDGFRB* (NM_002609.4):c.2586+5G>A variant. RNA-seq in proband G1-22 (red) demonstrates intron retention in contrast to normal splicing observed in unrelated inhouse control sample (blue). The aberrant transcript (r.2586_2587ins2586+1_2587-1) is predicted to result in p.(Thr863Valfs*70).

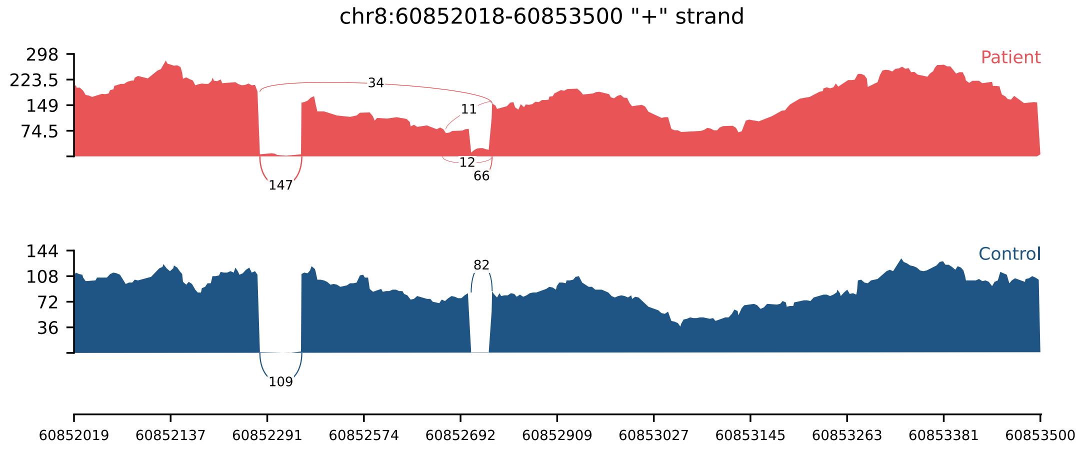


**Supplementary Figure S17:** Sashimi plot of heterozygous *CHD7* (NM_017780.4):c.6103+5G>T variant. RNA-seq in proband G1-24 (red) demonstrates exon skipping and 31-bp or 35-bp exon shortening due to alternative donor site usage in contrast to normal splicing observed in unrelated inhouse control sample (blue). The aberrant splicing produces three transcripts (r.[5895_6103del, 6069_6103del, 6073_6103del]), predicted to result in p.[(Trp1966Thrfs*9), (Val2025Asnfs*8), (Arg2024Thrfs*9)], respectively.

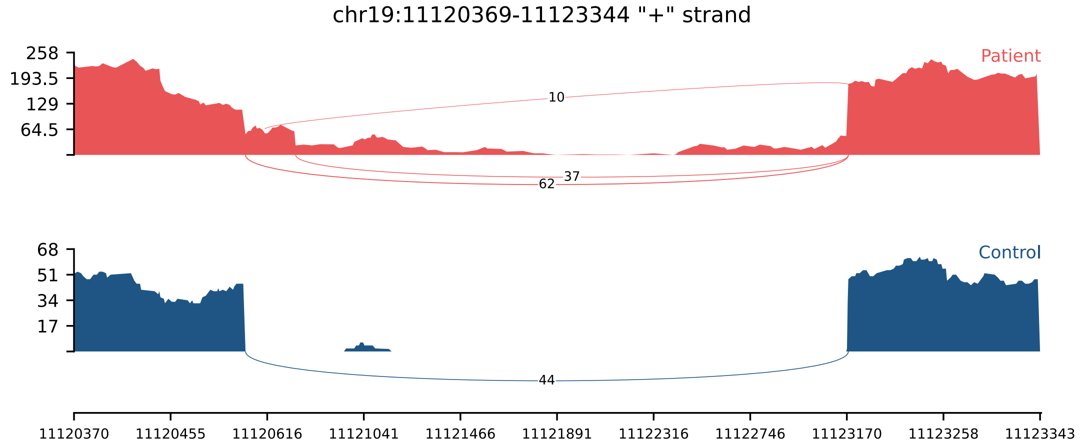


**Supplementary Figure S18:** Sashimi plot of heterozygous *LDLR* (NM_000527.5):c.2139A>G(p.Thr713=) variant. RNA-seq in proband G1-25 (red) demonstrates 217-bp or 81-bp exon extension due to alternative donor site usage, and intron retention in contrast to normal splicing observed in unrelated inhouse control sample (blue). The aberrant splicing produces three transcripts (r.[2140_2141ins2140+1_2140+217, 2140_2141ins2140+1_2141-1, 2140_2141ins2140+1_2140+81]), predicted to result in p.[(Glu714Glyfs*12), (Glu714Glyfs*12), (Glu714delinsXaa[28])], respectively.

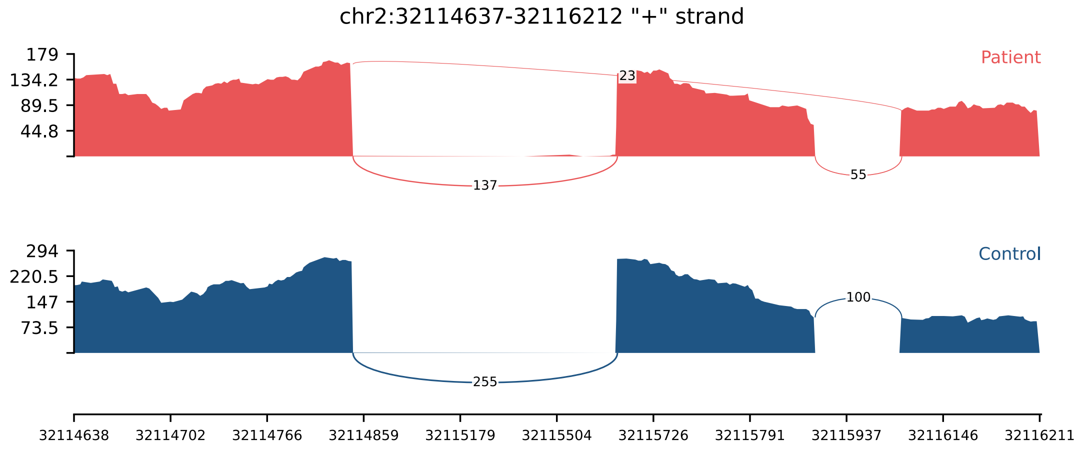


**Supplementary Figure S19:** Sashimi plot of heterozygous *SPAST* (NM_014946.4):c.1004+5G>A variant. RNA-seq in proband G1-26 (red) demonstrates exon skipping in contrast to normal splicing observed in unrelated inhouse control sample (blue). The aberrant transcript (r.871_1004del) is predicted to result in p.(Gly291Trpfs*5).

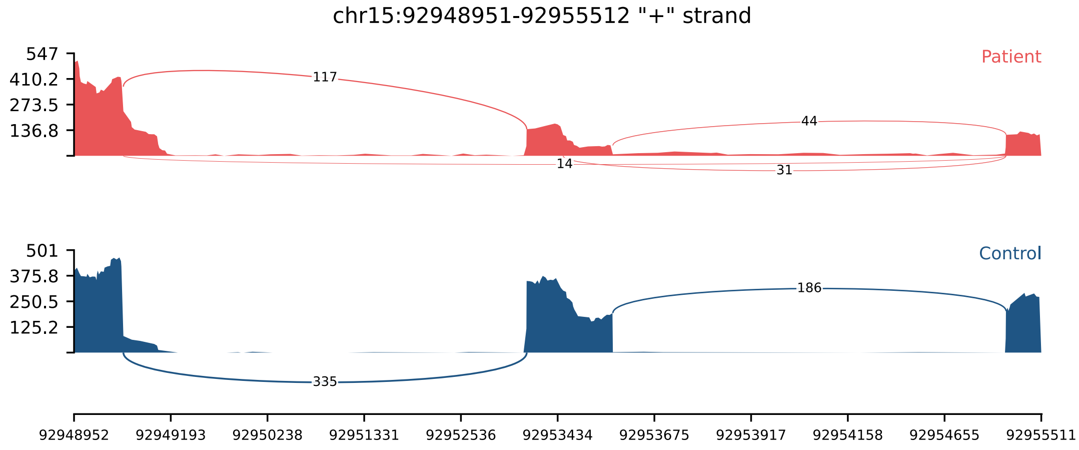


**Supplementary Figure S20:** Sashimi plot of heterozygous *CHD2* (NM_001271.4):c.1719G>A(p.Thr573=) variant. RNA-seq in proband G1-27 (red) demonstrates 124-bp exon shortening due to alternative donor site usage and exon skipping in contrast to normal splicing observed in unrelated inhouse control sample (blue). The aberrant splicing produces two transcripts (r.[1596_1719del, 1503_1719del]), predicted to result in p.[(Pro535Asnfs*12), (Asn502Tyrfs*14)], respectively.

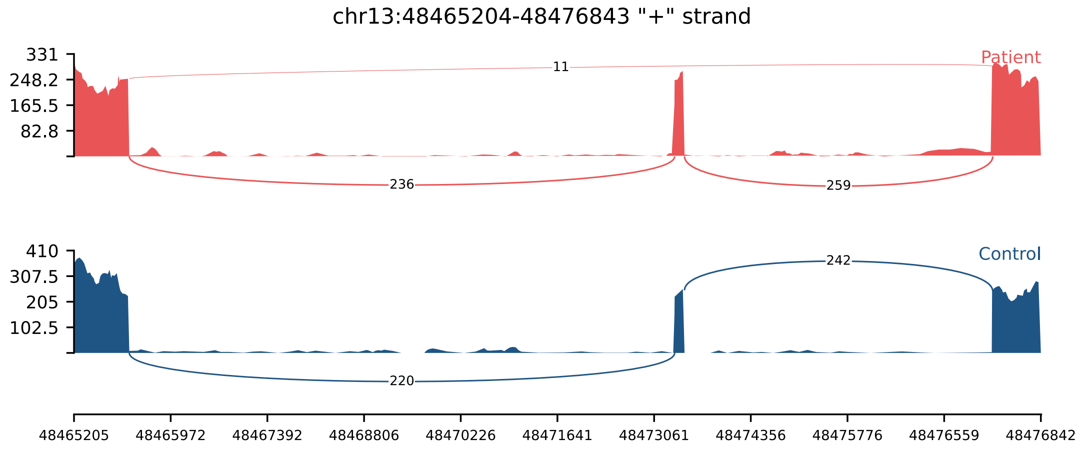


**Supplementary Figure S21:** Sashimi plot of heterozygous *RB1* (NM_000321.3):c.2520+5G>A variant. RNA-seq in proband G1-30 (red) demonstrates exon skipping in contrast to normal splicing observed in unrelated inhouse control sample (blue). The aberrant transcript (r.2490_2520del) is predicted to result in p.(Ile831Leufs*8).

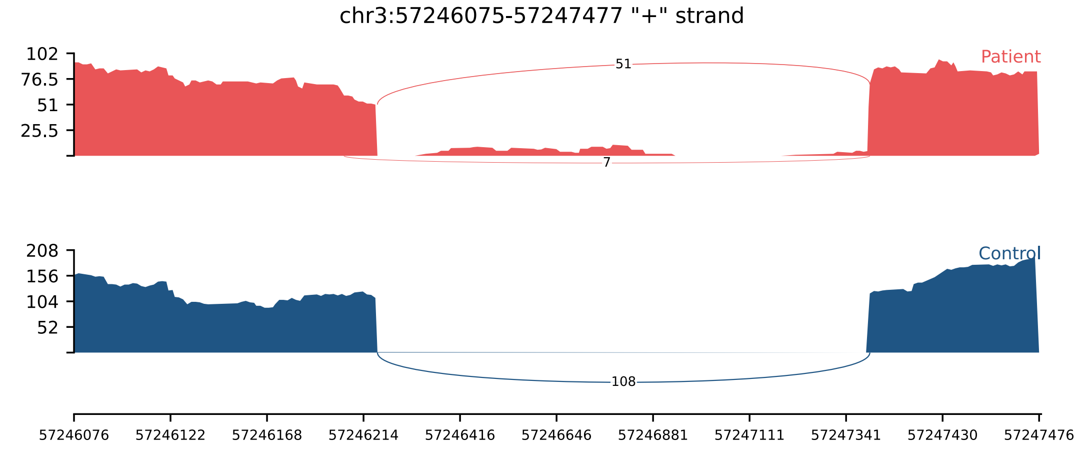


**Supplementary Figure S22:** Sashimi plot of heterozygous *APPL1* (NM_012096.3):c.621G>C (p.Gln207His) variant. RNA-seq in proband G2-7 (red) demonstrates 16-bp exon shortening due to alternative donor site usage in contrast to normal splicing observed in unrelated inhouse control sample (blue). The aberrant transcript (r.606_621del) is predicted to result in p.(Gly202Glufs*2).

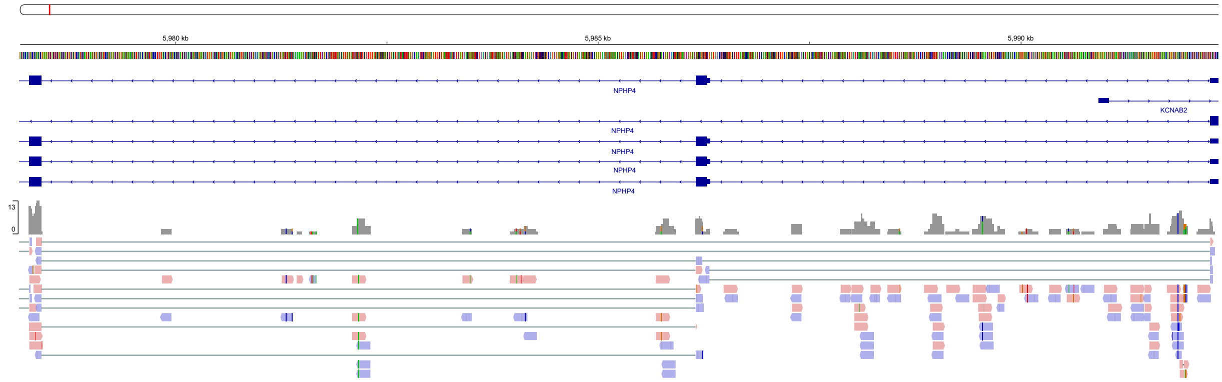


**Supplementary Figure S23:** IGV of the homozygous *NPHP4* (NM_015102):c.-38-10_-31del variant illustrating insufficient read coverage at the candidate locus, with only five reads spanning exons 1–2, one of which indicated alternative acceptor site usage and two of which indicated exon skipping. The *NPHP4* expression z-score in this proband was −1.54, and neither DASPER nor FRASER produced a statistically significant score for this region. Owing to the limited read depth, the consequence of the variant could not be reliably assessed, and it therefore remained classified as a variant of uncertain significance.

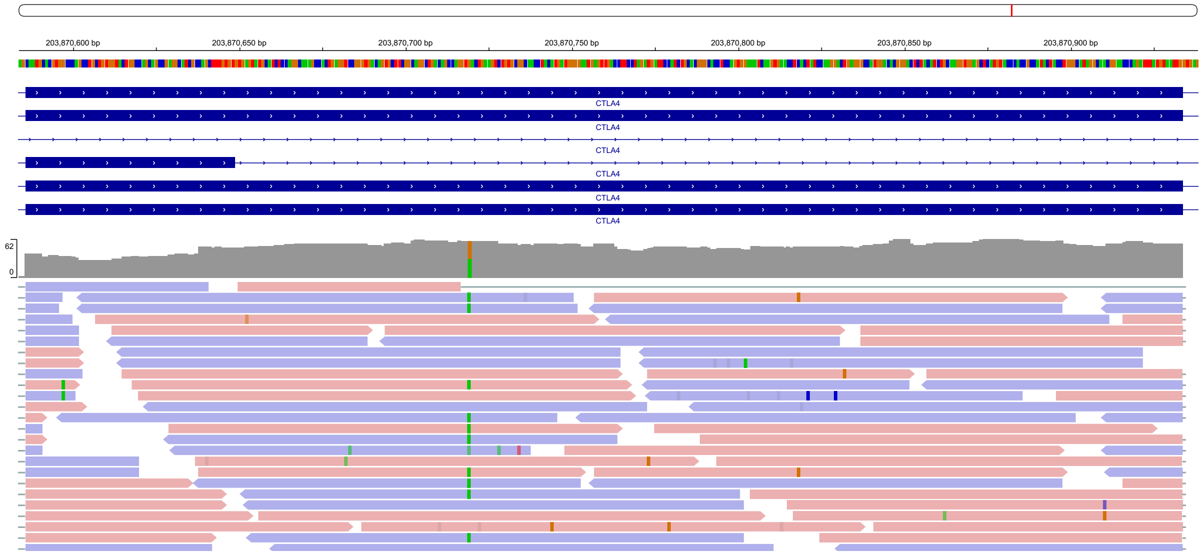


**Supplementary Figure S24:** IGV of the heterozygous *CTLA4* (NM_005214.5):c.243G>A (p.Val81=) variant illustrating an aberrant splicing event supported by minimal read evidence. The variant showed SpliceAI-predicted alternative donor site usage (217-bp exon shortening) in only a single read, whereas the remaining reads spanning the junction supported canonical donor site usage. The *CTLA4* expression z-score was −1.62; dasper produced an IFM score of 7753, whereas FRASER did not generate a score for this variant.

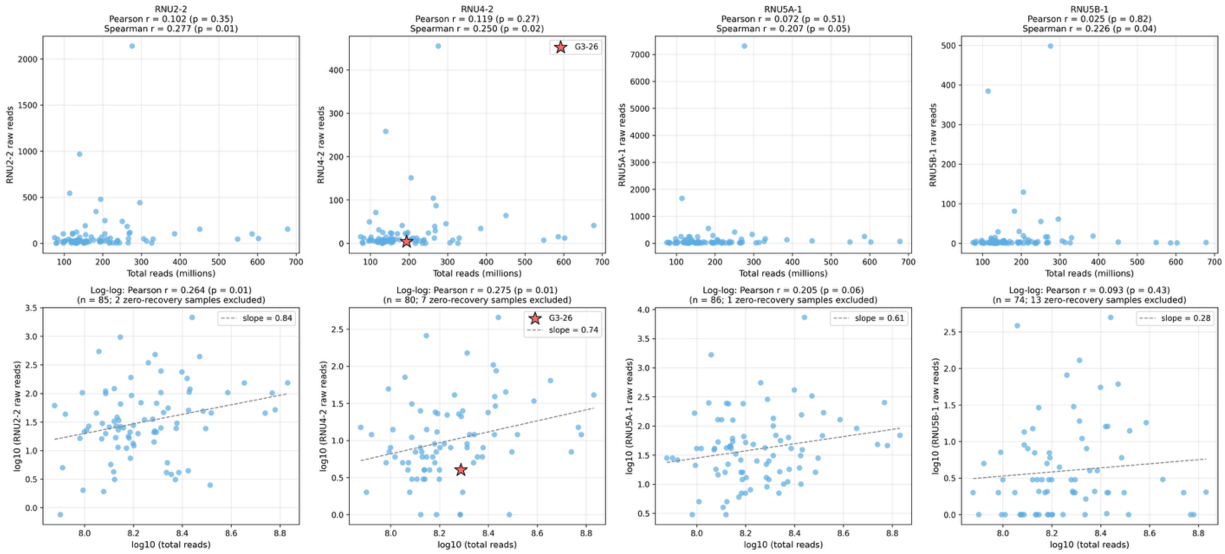


**Supplementary Figure S25:** Cohort-level analysis of the relationship between sequencing depth and snRNA gene recovery across the 87 QC-passing samples. Scatter plots of total post-trimming read count versus raw read counts assigned to the four snRNA genes (*RNU2-2*, *RNU4-2*, *RNU5A-1*, *RNU5B-1*). The top row shows all 87 samples on a linear scale, including those with zero snRNA recovery, with Pearson and Spearman correlation statistics; the bottom row shows the same relationship on a log10-transformed scale restricted to samples with detectable expression (sample number indicated per panel), as logarithmic transformation is undefined for zero values, together with the slope of the linear regression fit. On the log-log scale, snRNA recovery scales sub-linearly with library size across all four loci (slopes 0.28–0.84), indicating that library size contributes to but does not proportionally determine snRNA recovery. The *RNU4-2*–positive case G3-26 is highlighted (red star) and shows a below-median *RNU4-2* TPM (1.8; 23rd percentile of the cohort), indicating that adequate variant detection was achieved despite modest overall *RNU4-2* recovery at this locus.
